# Estimating the effective reproduction number from wastewater (R_t_): A methods comparison

**DOI:** 10.1101/2024.11.06.24316856

**Authors:** Dustin T. Hill, Yifan Zhu, Christopher Dunham, Joe Moran, Yiquan Zhou, Mary B. Collins, Brittany L. Kmush, David A. Larsen

## Abstract

**Background:** The effective reproduction number (R_t_) is a dynamic indicator of current disease spread risk. Wastewater measurements of viral concentrations are known to correlate with clinical measures of diseases and have been incorporated into methods for estimating the R_t_.

**Methods:** We review wastewater-based methods to estimate the R_t_ for SARS-CoV-2 based on similarity to the reference case-based R_t_, ease of use, and computational requirements. Using wastewater data collected between August 1, 2022 and February 20, 2024 from 200 wastewater treatment plants across New York State, we fit eight wastewater R_t_ models identified from the literature. Each model is compared to the R_t_ estimated from case data for New York at the sewershed (wastewater treatment plant catchment area), county, and state levels.

**Results:** We find a high degree of similarity across all eight methods despite differences in model parameters and approach. Further, two methods based on the common measures of percent change and linear fit reproduced the R_t_ from case data very well and a GLM accurately predicted case data. Model output varied between spatial scales with some models more closely estimating sewershed R_t_ values than county R_t_ values. Similarity to clinical models was also highly correlated with the proportion of the population served by sewer in the surveilled communities (r = 0.77).

**Conclusions:** While not all methods that estimate R_t_ from wastewater produce the same results, they all provide a way to incorporate wastewater concentration data into epidemic modeling. Our results show that straightforward measures like the percent change can produce similar results of more complex models. Based on the results, researchers and public health officials can select the method that is best for their situation.

**Key messages:**

1. Wastewater data has been used to estimate the R_t_ in different ways but the relative strengths and weaknesses of each method were unknown.
2. R_t_ estimation results from wastewater data are influenced by sewershed population size and geographic aggregation making selection of the best method dependent on the study location and available data.
3. Estimating the R_t_ from wastewater is desirable because wastewater data are anonymous, comprehensive, and efficient for measuring disease burden.

## 1 Introduction

Following an initial outbreak, the effective reproduction number or R_t_, quantifies secondary infections caused by an individual at a given time (t), and is a dynamic indicator of current disease spread risk (1). R_t_ provides an understanding of the effectiveness of public health interventions to counter transmission, behavioral adaptations of the population affected, the development of immunity in the population affected, and variant-specific characteristics (2). If an outbreak proceeds to establish endemic transmission, R_t_ becomes a timely and context-specific decision-influencing metric for public health officials.

R_t_ can be estimated different ways including from mechanistic (3) and statistical models (4) that are built using data from clinically reported cases. Cori et al. (5) proposed a method for calculating the R_t_ that is flexible and allows users to easily estimate R_t_ for a given pathogen with an open-source R package called EpiEstim (6). Cori et al.’s (5) method estimates R_t_ using the ratio of new infections to the total infectiousness of infected individuals at a specific time step. Their method is simple to implement and has become a common way to calculate the R_t_ for a given pathogen with many researchers using this method (2,7,8). Expanding beyond clinical surveillance data to other data inputs for estimating the R_t_ is a new direction for epidemiology and wastewater surveillance has emerged as a novel data input for epidemic models (9,10).

Wastewater surveillance data collected from communities has been an important tool for tracking infectious diseases (11) and is not limited by biases in case reporting that might result from lack of medical services or asymptomatic spread (12). The levels of SARS-CoV-2 RNA in wastewater have been shown to be a leading indicator of clinical COVID-19 cases (13–15).

Estimating R_t_ through wastewater-based epidemiology (WBE) presents an innovative approach that leverages a population-level indicator of infection prevalence (16). By integrating wastewater data into estimates of total infections, researchers can infer the temporal trends and changes in R_t_ (13,14). The growth of wastewater surveillance since the COVID-19 pandemic has led to a number of studies that propose different methods for transforming wastewater concentration values into estimations of R_t_.

Methods for estimating the R_t_ from wastewater fall into three general approaches. The first approach uses a model to predict cases from wastewater and then use the predicted cases to estimate R_t_ (9,17). The second approach involves a deconvolution of case data as determined by wastewater concentration estimates such as by dividing all wastewater concentrations collected for a time interval by the lowest detection ever reported and assuming that this low detection value represents a single case (10). These deconvoluted case data are then used in estimating the R_t_ (10,18,19). The third approach is to transform wastewater concentration data into an R_t_ value from some other model with researchers proposing the use of SEIR models (20–22) or an artificial neural network model (23). Given the potential utility of wastewater data as a measure of overall infections and the growing number of approaches to using WBE data to calculate R_t_, it is necessary to better understand the advantages and disadvantages of the various methods.

Using COVID-19 as our example, we compare eight unique methods of estimating the R_t_ from wastewater (five gathered from the literature and three proposed in this publication) to an R_t_ estimate from case data. We use all included methods for each approach to calculate R_t_ values for COVID-19 using SARS-CoV-2 wastewater data collected in New York State, USA and then compare each method to the R_t_ from case data.

## 2 Methods

### 2.1 Data collection

SARS-CoV-2 viral concentrations were calculated from wastewater samples collected from 205 wastewater treatment plants (WWTPs) across all 62 NYS counties for samples collected between August 1, 2022 and February 20, 2024. Five different laboratories across the state test the wastewater for SARS-CoV-2, each using a different laboratory method. Complete descriptions of the extraction and quantification methods have been previously described (24) and is included in the supplementary material. These data are publicly available from NYS Department of Health (25).

County level COVID-19 case data including the number of tests conducted and number of tests positive were obtained from New York State DOH from their COVID-19 public data repository (26). Data were summed across the state to obtain statewide incidence.

### 2.2 Methods to estimate R_t_

We conducted a search of preprint and peer-reviewed literature that utilized wastewater-based epidemiology in the calculation of the R_t_ between the dates of March 1, 2020 and July 30, 2024. We identified fourteen publications that met these criteria. We then selected methods from papers that were fully reproducible and written in the R coding language. This yielded five distinct methodologies across the publications. Three of these methods are published R packages including ERN (effective reproduction number) (27) (GitHub https://github.com/phac-nml-phrsd/ern), EpiSewer (28) (GitHub https://github.com/adrian-lison/EpiSewer), and ConcRt (20) (GitHub https://github.com/igoldsteinh/concRt). Two methods were not put into R packages, but have published R scripts in their supplemental materials (9,10,17). We include three additional R_t_ estimation methods that were derived by our research team (see Table 1 for the complete list of eight methods we explored along with equations).

**Table 1:** R_t_ estimation methods, citations, and equations.

| Method | Citation | Explanation | Equations<sup>†</sup> |
| --- | --- | --- | --- |
| Reference method ($R_t$ from case data) | Cori et al. (5) | EpiEstim method that estimates the $R_t$ as a function of the number of incident cases divided by the sum of cases during the serial interval. | $R_t = \frac{I_t}{\sum(I_{t-s}) * w_s}$ |
| Method 1 – Fit line | Proposed in the current manuscript – based upon commonly used estimates of the trend in levels of SARS-CoV-2 RNA in wastewater | $R_t$ is calculated from case data using the EpiEstim method for a given interval. (i.e., 45 days), then the wastewater quantification level on the same days is regressed against the $R_t$ value to get a slope and intercept for transforming viral RNA copies into a pseudo $R_t$ value ( $R_{t\ new}$ ). | $R_t = \frac{I_t}{\sum(I_{t-s}) * w_s}$ $i = 1 \text{ to } 45$ $t \text{ is any given day}$ $R_{ti} \sim \beta_1 * G_{ti} + \beta_0$ $R_{t \text{ new}} = \widehat{\beta}_1 * G_{t \text{ new}} + \widehat{\beta}_0$ |
| Method 2 – Rolling GLM | Daza-Torres et al.(9) and Montesinos-López et al.(17) published theoretically similar approaches, but used Bayesian frameworks for their case predictions instead of a GLM. | Cases are predicted from viral copies using a GLM. Wastewater predicted cases ($C_t$) are then placed in the EpiEstim equation. | $C_t = \sim poisson(Beta0, Beta, error)$ $R_t = \frac{C_t}{\sum C_{t-s}) * w_s}$ |
| Method 3 – EpiEstim substitution | Proposed in the current manuscript | Direct substitution replaced the incidence value ($I_t$) with a wastewater value ($G_t$). | $R_t = \frac{G_t}{\sum (G_{t-s}) * w_s}$ |
| Method 4 – Exponentiated percent change rate | Proposed in the current manuscript – based on commonly used metrics for the interpretation of wastewater surveillance results | The percent change approach estimates $R_t$ as a relative change of wastewater concentration ($G_{t-0}$) over the previous time period. The value is then inverted to put it on the same scale as the $R_t$. | $R_t = \frac{1}{e^{abs(\frac{G_{t-0}}{G_{t-1}-G_{t-0}})}}$ |
| Method 5 – Huisman et al. | Huisman et al.(10) | Observed gene copy quantification levels are transformed into daily infection incidence with the shedding load distribution to get a synthetic case value ($C_i$) that is used in the EpiEstim method. | $C_i = N * M \sum_j w_i - {}_j I_j$ $R_t = \frac{C_i}{\sum (C_{i-t-s}) * w_s}$ |
| Method 6 – Goldstein Et al. | Goldstein et al. (20) | An compartmental model is built and used to estimate the $R_t$ using four compartments: current observed infections (E) unobserved infections (I), recovered population (R1) and the starting $R_t$ value (R2). | E, I, R1, and R2 priors<br>$\frac{dE}{dt} = \alpha_t * I - \gamma * E$ $\frac{dI}{dt} = \gamma * E - v * I$ $\frac{dR1}{dt} = v * I - \eta * R1$ $\frac{dR2}{dt} = \eta * R1$ $R_{0,t} = \frac{\beta_t}{v}, R_t = R_{0,t} * \frac{S(t)}{N}$ |
| Method 7 – EpiSewer | Lison et al. (29) | Realized infections ($I_{t-s}$) are compared to reported wastewater concentration values and flow data to estimate a load-per-case ($u^{load}$) that is used for site specific estimation of the $R_t$ that is then smoothed with a spline approach. | $i_t = E[I_t] = R_t \sum_{s=1}^G \tau_s^{gen} I_{t-s} \vert t > G$ $\omega_t = \sum_{s=0}^s \zeta_{t-s} u^{load} \tau_s^{shed}$ |
| Method 8 – effective reproduction number R package (ERN) | Champredon et al. (27) | ERN transforms wastewater cases in a similar matter to Huisman et al. (10) with the addition of a LOESS smooth. The deconvolved case data are then input into the EpiEstim equation. | $w_t = \omega \sum_{k=1}^{i-1} i(t-k) f(k)$ $W_t = \text{smooth interpolation}(w_t, \theta)$ |
| Notes: † For method published by other authors, see citing publications for further notes and details on the equations. |  |  |  |

### 2.3 Estimation of R_t_

We applied the methods listed in Table 1 to the estimation of R_t_ at different spatial scales (statewide, county level, and sewershed level). Statewide wastewater concentration values were estimated using population weighted data from the sampling sites. Data were transformed into daily estimates from once and twice weekly sampling schedules using a two-stage approach.

First, data were linearly interpolated between sample days. Second, a 7-day moving average was calculated from the daily interpolation. Daily data were used as inputs for all eight models at the state level. For the county level analysis, we used the same population weighting approach aggregating up to the county using the daily interpolated values. Five counties were excluded because they had inconsistent reporting or large gaps in their time series leaving fifty-seven fifty-seven counties to analyze for all eight methods.

Last, for the sewershed level data, no population weighting was needed for transformation to daily data because the data were at the sampling site level. Sewershed case data were not available and county cases were used as a substitute in the comparison. All eight methods were applied to nine selected sewersheds from the two-hundred sampling sites in NYS. We did not produce estimates for all sewersheds because Method 7 required daily wastewater flow data, which was not available for most sites. To ensure a fair comparison for Method 7, the nine sewersheds selected all had the most consistent reporting of flow data limiting any bias based on method. Across all three analyses (state, county, and sewershed), six methods required an additional post-calculation step where the R_t_ values were smoothed using a 5-week moving average (centered) to reduce error and noise in the estimates.

### 2.4 Evaluation of model performance

Using guidance for comparing modeling approaches (30), we selected five metrics to evaluate accuracy when compared to the reference method (R_t_ from cases). First we calculated a value for percent agreement between the two R_t_ values based on if both statistics were above one at the same time or below one at the same time. This metric relies less on precision and magnitude of the change in R_t_ and more on whether both models report increases (above one) or decreases (below one) at the same time. Second, we calculated the root mean square error (RMSE) between the case R_t_ and each wastewater R_t_. Third, we compared the Pearson correlation between the case R_t_ and the wastewater R_t_. Fourth, we observed the time at which R_t_ values peaked in the data and determined if the wastewater R_t_ peaked in the same week. Fifth, we calculated the absolute difference between the case R_t_ and the wastewater R_t_ estimates. Sixth and finally, we calculated the sharpness of the R_t_ methods. Sharpness is the weighted average of the credible intervals across each estimated R_t_. We used seven coverages for the intervals (70%, 75%, 80%, 85%, 90%, 95%, 99%) with weights of 1 x (1 – coverage%). (30).

### 2.5 Influence of population coverage

Wastewater surveillance data may be influenced by variation in population coverage and sampling frequency. To test for this, we calculated separate R_t_ values for each county in NY and estimated separate model performance measures (e.g., RMSE). Then, we used the following equation to evaluate accuracy between the methods across the counties: accuracy measure ∼ sampling frequency + number of sampling sites + log (total population on sewer)

Model accuracy might be influenced by factors associated with WBE methods including sampling frequency, the number of sampling sites in a county, and the number of people covered by the surveillance system in the county.

## 3 Results

### 3.1 R_t_ model performance

Between September 2022 and February 2024, NYS experienced three distinct rises in new COVID-19 cases and each rise was correlated with a rise in wastewater detection of SARS-CoV- 2 (Figure 1). The R_t_ when calculated from new COVID-19 cases, peaked prior to peaks in case data and crossed the threshold of 1 during the week when new case data peaked (Figure 1).

**Figure 1:**
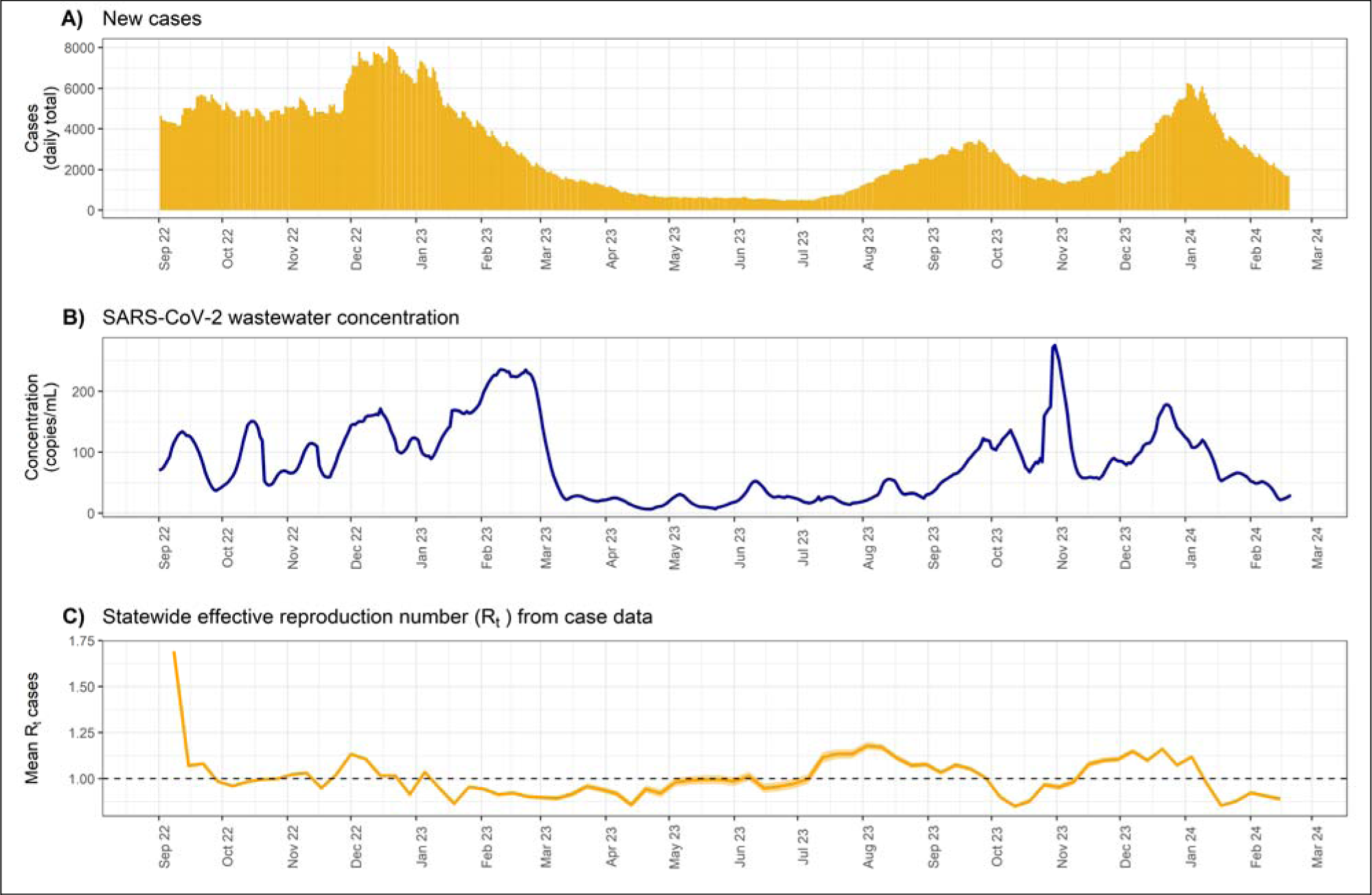
A) NYS new COVID-19 cases. B) NYS SARS-CoV-2 wastewater concentration. C) NYS Rt calculated from case data. Hashed lines denote peaks in new case data.

Model output generally followed the same trend as the R_t_ from case data (Figure 2). All methods identified the rise in case data from November 2023 to December 2023 as well as two prior rises in May 2023 and then in August 2023, corresponding with increases in the amount of SARS-CoV-2 in wastewater during those months and an increase in case reporting in August 2023 (Figures 1 and 2).

**Figure 2:**
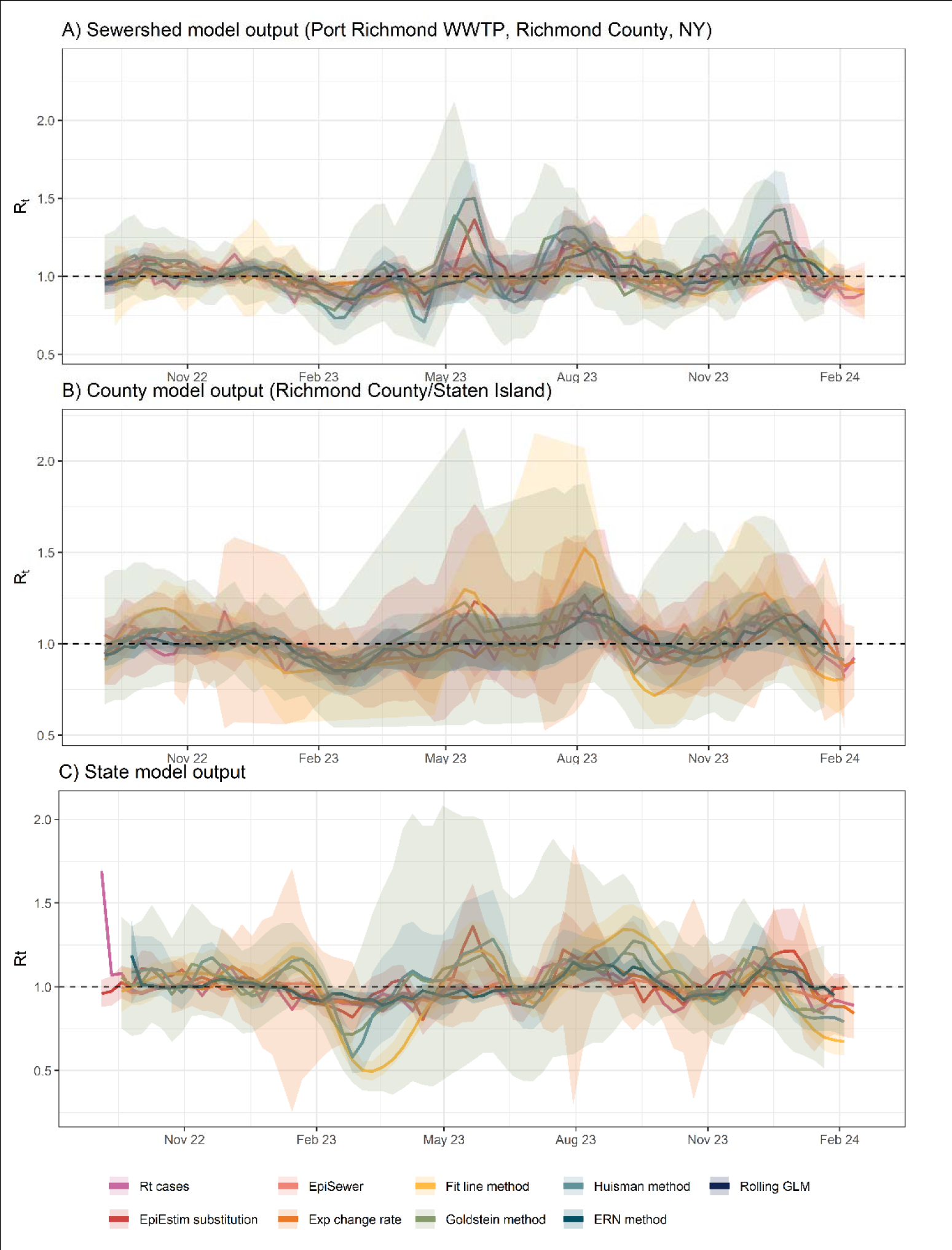
A) R_t_ for Port Richmond WWTP (Goldstein method excluded for exceptionally wide confidence intervals. B) R_t_ for Richmond County (Exp change rate method excluded for exceptionally wide confidence intervals). C) R_t_ for NYS (EpiSewer method excluded for exceptionally wide confidence interval).

Each R_t_ model required different input data to produce results and varied in their speed for calculating the R_t_. The Fit line method, the Rolling GLM, the Goldstein method, and EpiSewer method all take case data in addition to wastewater data as parameters (Table 2). The EpiEstim substitution method as well as the Exponentiated change rate, and the Huisman method do not require case data, and the ERN method data has an option to include case data, but it is not required (Table 2). The other key parameter that is required is flow data, but only the EpiSewer model requires daily reported flow data with no missing values, although imputation can be used to fill missing observations. Most methods were able to produce output in less than 1 minute for standardized datasets containing 93 observations (one sewershed sampled weekly for 3 months with weekly data transformed into daily data using interpolation), however, two methods took longer. The Goldstein method took 1.33 hours for produce output for statewide model and 61 minutes for the sewershed model, and EpiSewer took 34.41 minutes to produce ouptput for the statewide model but only 2.48 minutes for the sewershed model (Table 2). Also, most of the methods required an additional step to smooth the model output to reduce the variance of the predicted R_t_ values. A rolling, centered, 5-week average was found to adequately smooth the data and reduce the noise to produce output that more closely resembled the R_t_ from case data.

**Table 2:** Model parameters, required data, and computation time for estimating the R_t_ for the different methods.

| Method | Case data | Other parameters | Computation time for statewide data* | Computation time sewershed data* | Extra smoothing step |
| --- | --- | --- | --- | --- | --- |
| 1) Fit line method | Yes | Time window (days) | 0.53646088 secs | 1.457639 secs | No |
| 2) Rolling GLM | Yes | Time window (days) | 5.80379891 secs | 6.283749 secs | Yes, 5 weeks |
| 3) EpiEstim Substitution | No | Optional serial interval | 5.48190403 secs | 5.256281 secs | Yes, 5 weeks |
| 4) Exp. Change rate | No | Time window (days) | 0.066 secs | 0.01973436 secs | No |
| 5) Huisman | No | Optional serial interval | 9.52 secs | 7.911776 secs | Yes, 5 weeks |
| 6) Goldstein | Yes | E,I,R priors<br>E = sum of cases 11 days prior to start date * 5 * 1/3<br>I = sum of cases 11 days prior to the start date * 5 * 2/3<br>R = starting $R_t$ calculated from case data 11 days prior to start date | 1.33 hours | 61 minutes | Yes, 5 weeks |
| 7) EpiSewer | Yes | Daily flow data or average flow | 34.41 minutes | 2.48 | Yes, 5 weeks for the county data, not necessary for the sewershed data |
| 8) ERN | Optional | LOESS span -> specified through length of dataframe (< 200 obs ~ 0.2, 200-500 ~0.15, > 500 ~ 0.1) | 1.34 secs | 0.9020097 secs | Yes, 5 weeks |

Notes: \*A 93 day dataset was used with 3 months of observations between, Nov 1, 2023-Feb 1, 2024.

### 3.2 Spatial variation

Each method varied in its ability to produce R_t_ estimates across spatial scales. We found that most wastewater methods were able to produce statewide estimates close to clinical models, however, the EpiSewer method did not perform very well at the state level and instead was most similar to the case-based model at the sewershed level (Table 3). The method with the most consistent performance across geographic scales was the Rolling GLM (Table 3).

**Table 3:** Evaluation of R_t_ methods. County values are the median result from 57 counties and sewershed values are the median result from 9 sewersheds. All methods were compared to the weekly R_t_ from case data for the same scale except sewersheds, which used the countywide R_t_ value for the sewershed.

|  |  |  |  |  |  |  |  |
| --- | --- | --- | --- | --- | --- | --- | --- |
| *model* | *scale* | *RMSE* | *Pearson Correlation* | *Percent of peaks that coincide* | *Above or below 1 percent agreement* | *Mean absolute difference* | *Sharpness* |
| 1)Fit line method | State♦ | **0.0550**† | **0.7846**† | 0.1290 | **0.8026**† | **0.0426**† | 2.8193 |
|  | County | 0.1723 | 0.3720 | 0.1000 | 0.4167 | 0.1275 | **2.6403** |
|  | Sewershed | 0.1760 | 0.6246 | **0.2500**† | 0.7692 | 0.1132 | 4.0907 |
| 2)Rolling GLM | State♦ | **0.0625** | **0.6856** | **0.2143**† | **0.7895** | **0.0491** | **0.0029**† |
|  | County | 0.1600† | 0.4106† | 0.1220 | 0.6132† | 0.1154† | 0.0131† |
|  | Sewershed | 0.0934 | 0.2562 | 0.0000 | 0.6923 | 0.0794 | 0.0162† |
| 3)EpiEstim Substitution | State | 0.1044 | 0.3653 | 0.0625 | 0.5000 | 0.0888 | **0.0091** |
|  | County | 0.2282 | 0.2075 | **0.1400** | **0.5769** | 0.1752 | 0.0166 |
|  | Sewershed♦ | **0.0753**† | **0.7619**† | 0.0000 | 0.5385 | **0.0582**† | 0.0341 |
| 4)Exp change rate | State♦ | **0.0758** | **0.4750** | 0.1290 | **0.6711** | **0.0581** | 0.0344 |
|  | County | 0.2162 | 0.2473 | **0.1463** | 0.5642 | 0.1529 | 0.0933 |
|  | Sewershed | 0.1257 | 0.4029 | 0.0000 | 0.3846 | 0.1023 | **0.0295** |
| 5)Huisman method | State♦ | **0.1128** | 0.3933 | 0.0667 | 0.5526 | **0.0928** | **0.7479** |
|  | County | 0.2460 | 0.2298 | **0.1220** | **0.5833** | 0.1820 | 0.9248 |
|  | Sewershed | 0.1202 | **0.5072** | 0.0000 | 0.4615 | 0.1055 | 1.0314 |
| 6)Goldstein - EIRR | State♦ | **0.1799** | **0.4574** | 0.0000 | 0.6053 | **0.1456** | **0.1999** |
|  | County | 0.2381 | 0.2834 | 0.0909 | 0.6075 | 0.1860 | 0.3669 |
|  | Sewershed | 0.4899 | 0.1835 | **0.1667** | **0.6154** | 0.4017 | 0.2710 |
| 7)Episewer | State | 0.4609 | 0.2215 | 0.1429 | 0.5921 | 0.3018 | 3.9021 |
|  | County | 0.2292 | 0.1324 | **0.1633**† | 0.5429 | 0.1753 | 2.9200 |
|  | Sewershed♦ | **0.2106** | **0.5192** | 0.0000 | **0.7692**† | **0.1338** | **2.4306** |
| 8)ERN method | State♦ | **0.1425** | **0.3844** | 0.1538 | 0.5132 | **0.1177** | **2.4093** |
|  | County | 0.4271 | 0.1516 | 0.0976 | **0.5849** | 0.3085 | 2.5992 |
|  | Sewershed | 0.5946 | 0.2382 | **0.2000** | 0.4615 | 0.4744 | 2.7093 |
| Notes: ♦ Identifies the geography that the $R_t$ method was closest to the case $R_t$ method. †Identifies the best score for that geography across all methods. **Bold** indicates the best score across the geographic scales for each method. | | | | | | | |

### 3.3 Model evaluation

Model performance varied across all 57 counties in NYS. Two models (Goldstein method and the Rolling gLM) were nearly identical in their agreement with the R_t_ from cases regarding their percent agreement for R_t_ values above or below 1 (Figure 3). The Rolling GLM also had the lowest RMSE, mean absolute difference, and one of the lowest sharpness scores (Figure 3). The Rolling GLM also had high Pearson correlation with the R_t_ from cases across all counties, however, the Rolling GLM did not have the best score for the percent of peaks that coincide.

**Figure 3:**
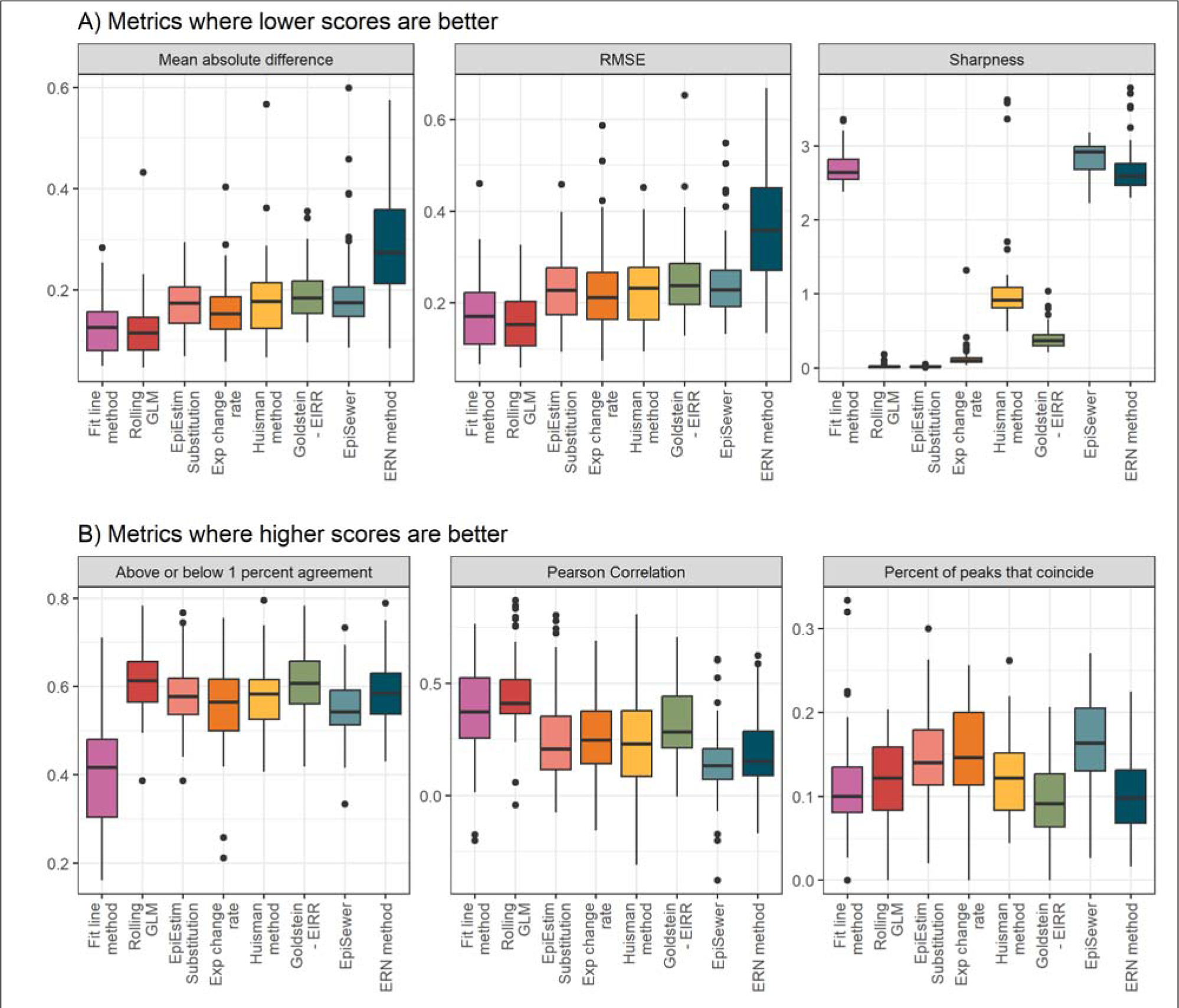
Evaluation metrics for all models across all 57 counties. A) Best scores are indicated by the lowest values. B) Best scores are indicated by the highest values.

Across all counties, the EpiSewer model had the highest percent agreement for peaks that occurred at the same time as the R_t_ from case data peaks (Figure 3).

Model outputs when compared to the R_t_ from case data correlated highly with the population served by the sewer system in each county. All methods reported positive correlations between r= 0.17 and r = 0.70 for the percent agreement and population on sewer in the county (Figure 4).

**Figure 4:**
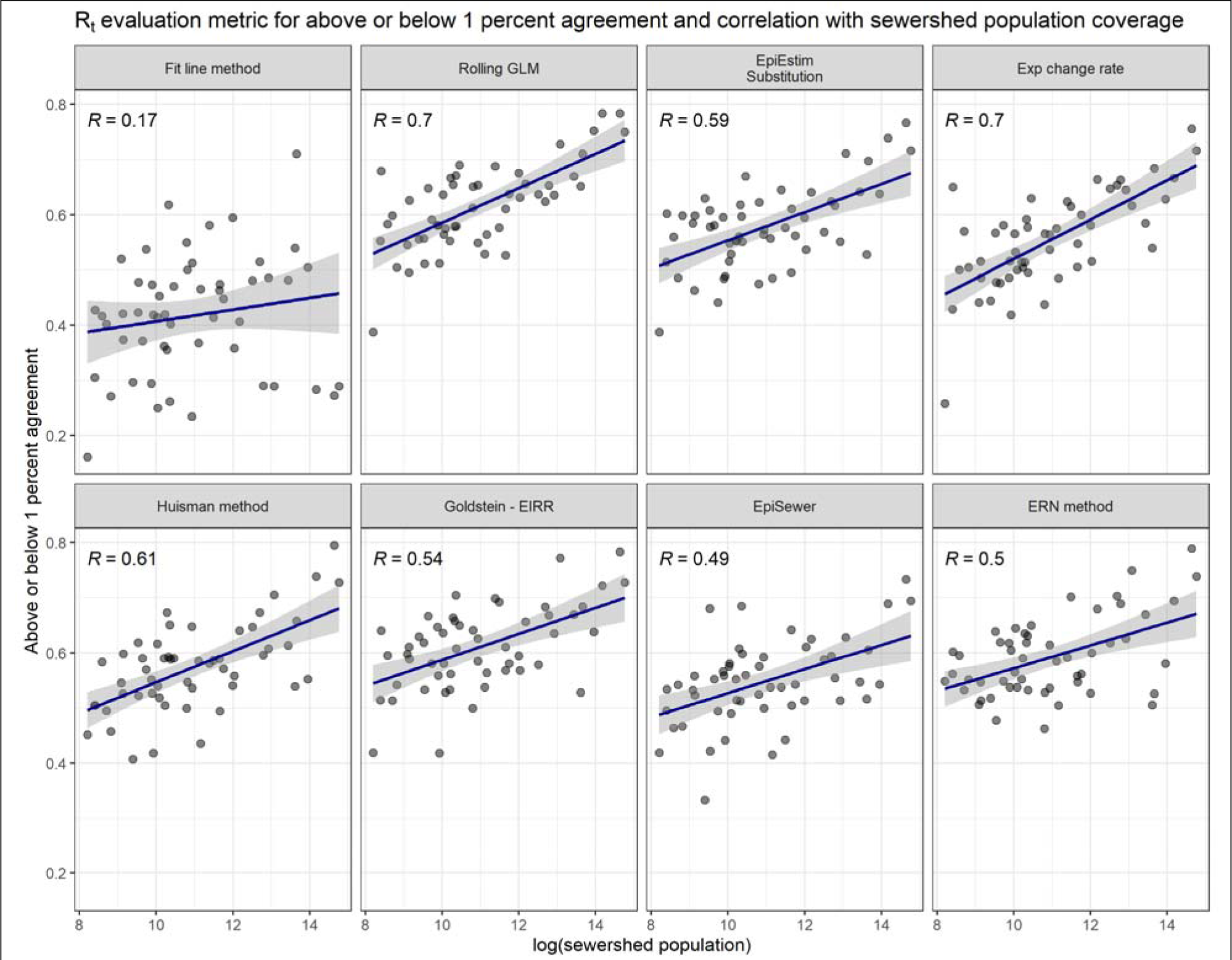
R_t_ evaluation results for the percent agreement for each model and correlation with population on sewer in each county.

In addition to population served, we also tested for predictive associations between sample frequency and the number of plants reporting wastewater data in each county. Across methods, we found that total population served was the most important predictor for percent agreement for values above or below 1 (Table 4). Three methods least impacted by population served were EpiSewer (β=0.012, *P* value = 0.143, Table 4), the fit line method (β=0.004, *P* value = 0.741, Table 4) and the Huisman method (β=0.014, *P* value = 0.060, Table 4). The only method that was moderately impacted by the number of WWTPs within the county was the fit line method (β=0.016, *P* value = 0.079, Table 4). All other methods were not significantly impacted by this factor. Sample frequency only moderately impacted one method, the Huisman method (β=0.071, *P* value = 0.098, Table 3) where greater sample frequency was associated with higher percent agreement.

**Table 4:** Regression results for R_t_ for agreement between the wastewater Rt and the case-based Rt for similar percent of values above or below one.

|  |  |  |  |  |  |  |  |  |
| --- | --- | --- | --- | --- | --- | --- | --- | --- |
|  | *1)Fit line method* | *2)Rolling GLM* | *3)EpiEstim substitution* | *4)Exp change rate* | *5)Huisman method* | *6)Goldstein method* | *7)EpiSewer* | *8)ERN method* |
| (Intercept) | 0.335<br>($<0.01^{***}$) | 0.321<br>($<0.01^{***}$) | 0.29<br>($<0.01^{***}$) | 0.33<br>($<0.01^{***}$) | 0.329<br>($<0.01^{***}$) | 0.33<br>($<0.01^{***}$) | 0.317<br>($<0.01^{***}$) | 0.361<br>($<0.01^{***}$) |
| log(sewershed population) | 0.004<br>(0.741) | 0.023<br>($<0.01^{***}$) | 0.022<br>($<0.01^{***}$) | 0.023<br>($<0.01^{***}$) | 0.014<br>(0.06•) | 0.023<br>($<0.01^{***}$) | 0.012<br>(0.143) | 0.017<br>(0.011**) |
| Number of WWTPs in the county | 0.016<br>(0.079•) | 0.002<br>(0.662) | -0.006<br>(0.222) | -0.004<br>(0.485) | 0.002<br>(0.776) | -0.004<br>(0.485) | 0.003<br>(0.657) | -0.006<br>(0.294) |
| Sample collection frequency (weekly) | -0.007<br>(0.92) | 0.03<br>(0.407) | 0.05<br>(0.178) | 0.025<br>(0.541) | 0.071<br>(0.098•) | 0.025<br>(0.541) | 0.067<br>(0.13) | 0.043<br>(0.286) |
| $R^2$ | 0.08 | 0.46 | 0.42 | 0.35 | 0.31 | 0.35 | 0.27 | 0.29 |
| n | 57 | 57 | 57 | 55 | 57 | 55 | 53 | 57 |
| Notes: *** $P$ value $< 0.01$, ** $P$ value $< 0.05$, • $P$ value $< 0.1$ | | | | | | | | |

## 4 Discussion

### 4.1 Methods for estimating R_t_ from wastewater

Multiple methods are available for the estimation of the R_t_ from wastewater data and when compared with the R_t_ from case data, methods that included clinical data were unsurprisingly closest to the case-based reference model. Of the eight methods reviewed, each has strengths and weaknesses. Interestingly, model accuracy was similar between methods with no method clearly performing above the others across all accuracy measures. Selection of a method might then be influenced by other factors like ease-of-use or whether there are available parameters like shedding rates and disease incidence. A decision tree with recommended method based on factors besides accuracy is provided in the supplementary material.

Further, two methods based on straightforward calculations of the GLM (method 2) of wastewater to case data (method 2) and percent change (method 4) yielded accurate results. This finding is important because it means that 1) additional modeling complexity might not yield much additional benefit, 2) the methods can most easily be adapted to any pathogen with quantifiable detection data, and 3) linear and generalized linear models can use wastewater to accurately predict new infections.

### 4.2 Spatial scale and R_t_ estimation

Spatial scale was found to be an important factor in R_t_ estimation and is something that public health agencies and researchers should consider when selecting a wastewater-based R_t_ method. Accuracy measures were higher at larger spatial scales (e.g., county was higher than sewershed), which follows a general observation in the literature that spatial aggregation yields increased correlations for wastewater and case data (31). Given the variability of wastewater concentration data and potential for less than daily sampling, at the sewershed level, we recommend the EpiEstim substitution method unless the data includes consistently reported flow data, then we recommend the EpiSewer method. Users that want to spatially aggregate the data to obtain estimates for wastewater concentration at larger scales should consider methods other than EpiSewer like the Rolling GLM.

### 4.3 Characteristics that influence the estimation of the R_t_

The total population served by the WWTP and the number of samplings sites in a community were associated with whether the model closely estimated the case-based R_t_ or not at the county level. Population on sewer was an influential variable for all methods except EpiSewer, Huisman, and the fit line method. Further, the number of sampling sites was influential for the fit line method only. This suggests that WBE methods for estimating the R_t_ should consider what portion of the community is covered by the sewer system so that R_t_ estimates can be interpreted or extrapolated at the county level. We did not find that sampling frequency influenced wastewater R_t_ estimates, which is important because increased sampling frequency can be costly and operationally difficult. The majority of sampling sites in NY sample once weekly and our results are suggestive that once weekly sampling is sufficient for R_t_ estimation from wastewater.

### 4.4 Applications beyond COVID-19

Of the methods reviewed, several would be applicable to other pathogens tested in wastewater, provided the tests provide quantifiable data for use in the R_t_ renewal equation. The exponentiated change rate and the EpiEstim substitution would be the simplest to adapt to a new pathogen given that they do not require any case data for the disease. The Huisman method is COVID-19 specific with the shedding rate for SARS-CoV-2 virus included in the equation, so adaptation to another pathogen would take extra work but it might be worthwhile since the method also does not require case data.

### 4.5 Limitations

One major limitation across models was the essential component for estimating the load to case ratio (EpiSewer, rolling GLM, ERN, and Goldstein) and this ties the models to the need for good reporting of new cases. Without assumptions for under-reporting, which the Goldstein method includes, each of these models will have the errors in case reporting reflected in the errors of their model output. Given that it is impossible to track all infections for COVID-19 this is an important limitation because of asymptomatic spread, but it might not be applicable to other diseases tracked in wastewater that are more readily reported. Lacking any better reference method, the case-estimated R_t_ value is still the best available model to compare wastewater-based R_t_ estimates. Additionally, for our sewershed level comparison, we did not have geocoded sewershed case data and only had county case data. This mismatch might have reduced the performance for the models at that scale.

## 5 Conclusion

Each research group and public health agency that collects wastewater data will have to decide the best R_t_ method for their program, and our findings provide a systematic way to select the optimal method given many different parameters. Selection of simpler models should not deter researchers since our results suggest they provide similar accuracy to more complex modeling approaches. Moving forward, the R_t_ as calculated by wastewater may become an important measure for established and emerging pathogens of concern.

## 6 Data availability

All data used in this study are publicly available from the New York State Department of Health website. Wastewater data are available at: https://health.data.ny.gov/Health/New-York-State-Statewide-COVID-19-Wastewater-Surve/hdxs-icuh/about_data, COVID-19 case data are available at: https://health.data.ny.gov/Health/New-York-State-Statewide-COVID-19-Testing/jvfi-ffup/about_data. In addition, aggregated data for NY counties, and statewide are included in the GitHub repository along with all code used for the analyses at https://github.com/dthill196/rt_wastewater.

## 7 Supplemental document

We provide a supplementary appendix with additional methods and sensitivity analyses.

## Supporting information

decision tree

## Author contributions

DTH, JM, MBC, BLK, and DAL designed the study and conceptualized the analysis. DTH, YZhou, JM, and BLK drafted the introduction and review of literature.

DTH, MBC, and DAL drafted the methods, results, and discussion. DTH, JM, BLK, MBC, YZhu, DAL Writing, review and editing

DTH, YZhu, JM, and YZhou conducted the data analysis and created the figures and tables.

DTH, CD, and YZhou developed scripts, software, and cloud computing resources for the project

DAL supervised the project.

## Funding

This work was supported by the CDC’s ELC Program, Award Number NU50CK000516. Computing resources were provided by Syracuse University HTC Campus Grid and NSF award ACI 1341006.

### Acknowledgements

The authors would like to thank Adrian Lison for assistance with the EpiSewer package, Volodymyr Minin and Isaac Goldstein for assistance with the ConcRt package and working in Julia. The authors also thank David Champredon at Public Health Agency of Canada for his help with the ERN package. The authors also thank the Syracuse University IT team including Daniel Jeski, Peter Pizzimenti, Larne Pekowsky, and Nick Neal for their assistance with cloud computing resources.

## Notes

### Competing Interest Statement

The authors have declared no competing interest.

### Funding Statement

This work was supported by the CDC ELC Program Award Number NU50CK000516. Computing resources were provided by Syracuse University HTC Campus Grid and NSF award ACI 1341006.

## References

1. Vegvari C, Abbott S, Ball F. Commentary on the use of the reproduction number R during the COVID-19 pandemic, 2022. Stat Methods Med Res. 2022;31(9):1675–85.

2. Gostic KM, McGough L, Baskerville EB, Abbott S, Joshi K, Tedijanto C, et al. Practical considerations for measuring the effective reproductive number, Rt. PLOS Comput Biol. 2020 Dec 10;16(12):e1008409.

3. Bettencourt LMA, Ribeiro RM. Real Time Bayesian Estimation of the Epidemic Potential of Emerging Infectious Diseases. PLOS ONE. 2008 May 14;3(5):e2185.

4. Wallinga J, Teunis P. Different Epidemic Curves for Severe Acute Respiratory Syndrome Reveal Similar Impacts of Control Measures. Am J Epidemiol. 2004 Sep 15;160(6):509–16.

5. Cori A, Ferguson NM, Fraser C, Cauchemez S. A New Framework and Software to Estimate Time-Varying Reproduction Numbers During Epidemics. Am J Epidemiol. 2013 Nov 1;178(9):1505–12.

6. Cori A. EpiEstim: Estimate Time Varying Reproduction Numbers from Epidemic Curves [Internet]. 2021. Available from: https://CRAN.R-project.org/package=EpiEstim

7. Estrada E. COVID-19 and SARS-CoV-2. Modeling the present, looking at the future. Phys Rep. 2020 Jul 10;869:1–51.

8. Sy KTL, White LF, Nichols BE. Population density and basic reproductive number of COVID-19 across United States counties. PLOS ONE. 2021 Apr 21;16(4):e0249271.

9. Daza-Torres ML, Montesinos-López JC, Kim M, Olson R, Bess CW, Rueda L, et al. Model training periods impact estimation of COVID-19 incidence from wastewater viral loads. Sci Total Environ. 2023 Feb 1;858:159680.

10. Huisman JS, Scire J, Caduff L, Fernandez -Cassi Xavier, Ganesanandamoorthy P, Kull A, et al. Wastewater-Based Estimation of the Effective Reproductive Number of SARS-CoV-2. Environ Health Perspect. 2022;130(5):057011.

11. Sims N, Kasprzyk-Hordern B. Future perspectives of wastewater-based epidemiology: Monitoring infectious disease spread and resistance to the community level. Environ Int. 2020 Jun 1;139:105689.

12. Peccia J, Zulli A, Brackney DE, Grubaugh ND, Kaplan EH, Casanovas-Massana A, et al. Measurement of SARS-CoV-2 RNA in wastewater tracks community infection dynamics. Nat Biotechnol. 2020 Oct;38(10):1164–7.

13. Olesen SW, Imakaev M, Duvallet C. Making waves: Defining the lead time of wastewater-based epidemiology for COVID-19. Water Res. 2021 Sep 1;202:117433.

14. Wu F, Lee WL, Chen H, Gu X, Chandra F, Armas F, et al. Making waves: Wastewater surveillance of SARS-CoV-2 in an endemic future. Water Res. 2022 Jul 1;219:118535.

15. Larsen DA, Wigginton KR. Tracking COVID-19 with wastewater. Nat Biotechnol. 2020 Oct;38(10):1151–3.

16. Kaplan EH, Zulli A, Sanchez M, Peccia J. Scaling SARS-CoV-2 Wastewater Concentrations to Population Estimates of Infection [Internet]. 2021 Jul [cited 2021 Dec 1] p. 2021.07.15.21260583. Available from: https://www.medrxiv.org/content/10.1101/2021.07.15.21260583v1

17. Montesinos-López JC, Daza-Torres ML, García YE, Herrera C, Bess CW, Bischel HN, et al. Bayesian sequential approach to monitor COVID-19 variants through test positivity rate from wastewater. mSystems. 2023 Jul 25;8(4):e00018–23.

18. Wannigama DL, Amarasiri M, Hongsing P, Hurst C, Modchang C, Chadsuthi S, et al. COVID-19 monitoring with sparse sampling of sewered and non-sewered wastewater in urban and rural communities. iScience. 2023 Jul 21;26(7):107019.

19. Amman F, Markt R, Endler L, Hupfauf S, Agerer B, Schedl A, et al. Viral variant-resolved wastewater surveillance of SARS-CoV-2 at national scale. Nat Biotechnol. 2022 Dec;40(12):1814–22.

20. Goldstein IH, Parker DM, Jiang S, Minin VM. Semiparametric inference of effective reproduction number dynamics from wastewater pathogen surveillance data. ArXiv. 2023 Aug 31;arXiv:2308.15770v2.

21. Nourbakhsh S, Fazil A, Li M, Mangat CS, Peterson SW, Daigle J, et al. A wastewater-based epidemic model for SARS-CoV-2 with application to three Canadian cities. Epidemics. 2022 Jun 1;39:100560.

22. Asadi M, Oloye FF, Xie Y, Cantin J, Challis JK, McPhedran KN, et al. A wastewater-based risk index for SARS-CoV-2 infections among three cities on the Canadian Prairie. Sci Total Environ. 2023 Jun 10;876:162800.

23. Jiang G, Wu J, Weidhaas J, Li X, Chen Y, Mueller J, et al. Artificial neural network-based estimation of COVID-19 case numbers and effective reproduction rate using wastewater-based epidemiology. Water Res. 2022 Jun 30;218:118451.

24. Hill DT, Alazawi MA, Moran EJ, Bennett LJ, Bradley I, Collins MB, et al. Wastewater surveillance provides 10-days forecasting of COVID-19 hospitalizations superior to cases and test positivity: A prediction study. Infect Dis Model. 2023 Dec 1;8(4):1138–50.

25. NYS DOH. New York State Statewide COVID-19 Wastewater Surveillance Data | State of New York [Internet]. [cited 2024 Aug 21]. Available from: https://health.data.ny.gov/Health/New-York-State-Statewide-COVID-19-Wastewater-Surve/hdxs-icuh/about_data

26. DOH. COVID-19 Data in New York | Department of Health [Internet]. 2022 [cited 2022 Dec 26]. Available from: https://coronavirus.health.ny.gov/covid-19-data-new-york

27. Champredon D, Papst I, Yusuf W. ern: An R package to estimate the effective reproduction number using clinical and wastewater surveillance data. PLOS ONE. 2024 Jun 21;19(6):e0305550.

28. Lison A, Julian TR, Stadler T. Improving inference in wastewater-based epidemiology by modelling the statistical features of digital PCR [Internet]. bioRxiv; 2024 [cited 2024 Oct 29]. p. 2024.10.14.618307. Available from: https://www.biorxiv.org/content/10.1101/2024.10.14.618307v1

29. Lison A. adrian-lison/EpiSewer [Internet]. 2024 [cited 2024 Aug 22]. Available from: https://github.com/adrian-lison/EpiSewer

30. Bracher J, Ray EL, Gneiting T, Reich NG. Evaluating epidemic forecasts in an interval format. PLOS Comput Biol. 2021 Feb 12;17(2):e1008618.

31. Zhu Y, Hill D, Zhou Y, Larsen DA. The Effect of the Modifiable Areal Unit Problem (Maup) on Spatial Aggregation of Covid-19 Wastewater Surveillance Data [Internet]. Rochester, NY: Social Science Research Network; 2024 [cited 2024 Oct 8]. Available from: https://papers.ssrn.com/abstract=4917915

