## supplementary material for "Estimating the effective reproduction number from wastewater (R_t_): A methods comparison"

Estimating the effective reproduction number from wastewater (R_t_): A methods comparison: Supplemental methods and analysis

**Authors**: Dustin T. Hill*, Yifan Zhu, Christopher Dunham, Joe Moran, Yiquan Zhou, Mary Collins, Brittany Kmush, and David Larsen

### 1 Supplemental methods

#### 1.1 Lab methods

Wastewater analysis methods have been reproduced from a previously published paper that used the same dataset (1). Wastewater samples were processed and analyzed for SARS-CoV-2 by four regional laboratories each with different methods (see supplemental Table 1 for brief descriptions).

| Supplemental table 1: Methods used by regional labs | | |
| --- | --- | --- |
| **Lab** | **Processing method** | **Quantification method** |
| NYC | Centrifuged the PEG precipitation | Reverse transcription quantitative polymerase chain reaction (RT-qPCR) |
| Quadrant | Ultracentrifugation through a sucrose cushion | Reverse transcription quantitative polymerase chain reaction (RT-qPCR) |
| Stony Brook | Centrifuged to remove debris prior to Polyethylene glycol (PEG) precipitation using Quagen QiAamp DSP viral RNA mini kit (Qiagen, Hilden, Germany) | Concentrations measured using digital PCR  NanoDrop One Spectrophotometer (Thermo Fisher Scientific, Waltham, MA.) |
| UB-SUNY | Nanotrap Magnetic beads | Reverse transcription quantitative polymerase chain reaction (RT-qPCR) |

##### 1.1.1 NYC Sample processing

Samples in New York City were processed by the NYC lab and consisted of twenty-four hour (24h) flow-weighted composite influent samples. Samples were transported on ice and stored at 4°C before being processed within twelve hours of collection. 40 mL aliquots of each 24h composite sample were pasteurized at 60°C for 90 minutes and then centrifuged to remove solids (5,000 x *g*, 4°C, 10 minutes). Then, the supernatant was filtered using 0.22 µm of cellulose acetate before being subjected to virus concentration using polyethylene glycol (PEG) precipitation. 4 g of PEG and 0.9 g of NaCl were used followed by overnight incubation at 4°C with centrifugation at 12,000 x *g* at 4°C for 120 minutes to pellet the viruses. Once complete, the supernatant was discarded and RNA and DNA were extracted from the concentrated PEG pellet using Qiagen QiAmp Viral RNA Mini Kit with modifications.

##### 1.1.2 NYC Quantification method

A real-time quantitative polymerase chain reaction (RT-qPCR) assay was used to quantify copies of the SARS-CoV-2 nucleocapsid or N gene targeting the N1 region in triplicate reactions on a StepOnePlus real-time PCR system from Thermo Fisher Scientific. Please see Hoar et al. 2022 for additional details.(2)

##### 1.1.3 Quadrant Sample processing

110 mL-1.9L 24h composite samples of influent wastewater were collected then stored at 4°C before being transported to the lab for processing and quantification. Wastewater samples were blended to resuspend particulates that had settled during transport or storage. 20 mL was transferred to an ultracentrifuge tube (Thermo Fisher, MA, USA). A 12 mL sucrose cushion was then added underneath the wastewater using a serological pipette keeping the wastewater and the sucrose solution as distinct layers in the ultracentrifuge tube. These were then ultracentrifuged at 150,000 x *g* at 4°C on a Sorvall® WX Ultra series with a Sorvall SureSpin® 630 Swinginig-Bucekt Rotor for 45 minutes (Thermo Fisher). The resulting pellets with the viral particles and nucleic acids were carefuly decanted with a new pipette and resuspended 200 µL 1X PBS and transferred to 1.7 mL microcentrifuge tubes. These resuspended pellets were stored at -20°C for less than twenty-four hours until nucleic extraction. Extraction was done using the AllPrep® PowerViral® DNA/RNA Kit from Qiagen (Hilden, Germany).

##### 1.1.4 Quadrant Quantification method

RT-qPCR was used to detect the presence of SARS-CoV-2 RNA in undiluted total nucleic extracts using a multiplex reaction with the IP2 and IP4 assays targeting separate regions of the RdRp gene. Thermal cycling for 10 minutes at 50°C, 10 minutes at 95°C, followed by 45 cycles of 95°C for 10 seconds and 59°C for 30 seconds was done. These methods have been previously published and described in detail by Wilder et al. 2022(3).

##### 1.1.5 Stony Brook Sample processing

24h composite samples of raw sewage were centrifuged at 4200 rpm for 30 min at 4°C to remove large particles and debris before polyethylene glycol (PEG) precipitation. Recovery rates were evaluated using bovine coronavirus (BCoV), which belongs to the same genus as SARS-CoV-2, was spiked into the supernatant. The viral particles in 40 mL of samples were precipitated with PEG 8000 (Millipore Sigma, Burlington, MA) and NaCl (5 M, Millipore Sigma, Burlington, MA) and then incubated overnight at 4 °C. RNA from the PEG-precipitated wastewater was extracted by Qiagen QIAamp DSP viral RNA mini kit (Qiagen, Hilden, Germany) according to manufacturer’s instructions and eluted in 100 µL by nuclease-free water. The concentrations of RNA were measured by NanoDrop One Spectrophotometer (Thermo Fisher Scientific, Waltham, MA). All RNA samples were stored at −80 °C and subjected to cDNA synthesis within the same day of RNA extraction to avoid losses associated with storing and freezing and thawing RNA extracts.

##### 1.1.6 Stony Brook Quantification method

Reverse transcription was performed by High Capacity RNA-to-cDNA Kit (Applied Biosystems, Waltham, MA) at 37 °C for 60 min, and stored at -20 °C until further analysis. The cycling condition was 95 °C for 10 min, followed by 40 cycles of 95 °C for 5 s and 55 °C for 40 s, and 98 °C for 10 min. The total volume of each reaction was 14.5 µL containing 7.25 µL of QuantStudio 3D Digital PCR Master mix v2 (Applied Biosystems, Massachusetts, USA), 0.725 µL of primer and probe (N1/ BCoV), 0.725 µL of TaqMan® Copy Number Reference Assay RNase P (as an internal control, Applied Biosystems, Waltham, MA), 4.8 µL of nuclease-free water, and 1 µL of cDNA template. Digital PCR was performed using N1 primers and probe set from 2019-nCoV CDC EUA Kit (IDT # 10006606) and BCoV set against the BCoV gene as an external reference on a QuantStudio 3D Digital PCR (Applied Biosystems, Massachusetts, USA). Nuclease-free water was used as non-template control (NTC) and plasmids containing the complete nucleocapsid gene from 2019-nCoV (IDT # 10006625) were used as a positive control. Data analysis was performed with the online version of the QuantStudio 3D AnalysisSuite Cloud Software.

##### 1.1.7 UB-SUNY Sample processing

Beginning April 18, 2022, processing method two for UB-SUNY samples took the 24h influent samples of 9.75 mL and mixed them with 100 µL of Nanotrap® Enhancement Reagent 1 (Ceres Nanosciences) and 150 µL of Nanotrap® Microbiome A Particles (Ceres Nanosciences). Viruses were separated from the wastewater using KingFisher Apex Benchtop Sample Prep system from Thermo Fisher. After separation, the nucleic acids were extracted using MagMAX Viral/Pathogen Nucleic Acid Isolation Kits (Thermo Fisher) then eluted in MagMAX Viral/Pathogen Elution Buffer (Thermo Fisher) and stored at -80°C.

##### 1.1.8 UB-SUNY Quantification method

Samples processed using method one and method two by the UB-SUNY lab were both quantified using the same procedures. UB-SUNY quantified SARS-CoV-2 N gene(4) using RT-qPCR. The RT-qPCR quantification used 10 µL RT-qPCR reaction mixtures consisting of 5 µL of 2x iTag Universal Probes Reaction Mix from Bio-Rad, 0.25 µL of 50x iScript reverse transcriptase also from Bio-Rad, 0.75 µL of 2019nCoV_N2 (RUO Kit, IDT), and 4µL of undiluted nucleic acid extracts. RT-qPCR of the nucleic acid extracts. The SARS-CoV-2 reactions were heated at 50°C for 15 minutes, 95°C for 1 minute, and 40 cycle of 95°C for 10 seconds and 60°C for 30 seconds. Each RT-qPCR assay was conducted in duplicates or triplicates on a CFX96 Touch Real-Time PCR Detection System (Bio-Rad).

#### 1.2 Rt methods

The full county weighting population was:

| $gene copies \left( pop weighted \right)= \frac{copies*sewershed population}{county population}$ | **Equation 1**: aggregating wastewater results to obtain a county estimate |
| --- | --- |

We provide detailed descriptions of each of the methods for each below. To estimate our comparator from case data, we use the equation published by Cori et al (5) (Equation 2). Using this equation required a serial interval mean and standard deviation for the time between cases and infections based on development of symptoms. We used the R EpiEstim (6) package to determine the serial interval using Markov Chain Monte Carlo (MCMC) to estimate the interval from the data to allow for more continuous calculation of the Rt over different waves of infection.

| $R_{t}= \frac{I_{t}}{\sum{(I}_{t-s})*w_{s}}$ | **Equation 2**: Cori et al. (5) estimated the *R_t_* as a function of the number of cases at time *t* (*I_t_*) divided by the sum of the number of cases during the serial interval (*I_t - s_*) multiplied by the generation interval (*w_s_*). The resulting Rt is an estimate for the reproductive number on day t based on the ratio of new incident cases compared to the previous t days new cases. The equation has been shown to be informative for estimating the Rt for COVID-19 (7). |
| --- | --- |

##### 1.2.1 Rt method 1: Fit line

The fit line method is built on the assumption that wastewater quantification levels of gene copies are linearly correlated with all active cases (8) . While this assumption has its limitations, including that some individuals shed more virus than others (9), it allows us to take the Rt estimated from case data and correlate it with raw wastewater data. The linear association between raw wastewater and the Rt from case data can then be used to transform wastewater gene copies into a number on the same scale as the Rt to produce an Rt from wastewater estimate using the following equations:

| $R_{t}= \frac{I_{t}}{\sum{(I}_{t-s})*w_{s}}$  $i=1 to 45$  $t is anygiven day$  $R_{ti} \sim\beta_{1}* G_{ti}+ \beta_{0}$  $R_{t new}=\hat{\beta_{1}}*G_{t new}+ \hat{\beta_{0}}$ | **Equation 3**: Using the EpiEstim method, calculate the $R_{t}$ from case data with a moving window of i values (i.e., 45 days) previous from the current time point. Determine the linear association between $R_{t}$and gene copies for the 45-day period. Then, extracting $\beta_{1}$ and $\beta_{0}$ for the 45-day interval, predict the value of $R_{t}$ from wastewater by using the current concentration of gene copies in wastewater. |
| --- | --- |

##### 1.2.2 Rt method 2: Rolling GLM

The rolling GLM method is partly informed by previously published methods from Montesino-López et al. (10) and Daza-Torres et al.(11) that propose estimating new cases from a separate model, with those authors using a Bayesian approach. We propose a modification that uses a simpler, generalized linear model to build a predictive model for cases from wastewater for a relevant rolling time window. This method maintains its theoretical similarity to the method used by Montesino-López et al.(10) and Daza-Torres et al.(11) without the Bayesian portion. The exact method proposed by these studies was unable to be directly reproduced because not data or code were provided with each publication.

Our rolling GLM method takes the following form:

| $C_{t}=\sim{poisson(Beta0, Beta, error)}$ | **Equation 4a:** Predict cases from wastewater using 45 days of data from time t |
| --- | --- |
| $R_{t}= \frac{C_{t}}{\sum C_{t-s})*w_{s}}$ | **Equation 4b**: use cases predicted by wastewater in the EpiEstim model (Equation 2) |

The coefficient for wastewater as a predictor of cases would then be multiplied by wastewater intensity or gene copy values (log transformed) to obtain estimates of case data for those days. Those case estimates would then be put into the EpiEstim equation to calculate the R_t_.

##### 1.2.3 Rt method 3: EpiEstim direct substitution

Using the Rt equation published by Cori et al. (2013), we estimated Rt from wastewater by substituting the incidence value for the normalized wastewater intensity or raw gene copies value. We assume that a wastewater result on day s is the estimated active infections in a community and new infections that are asymptomatic and will show up n days from infection based on the lag between shedding and symptoms. We followed the same structure as Equation [1] with some modification. The revised Rt from wastewater equation we used is:

| $R_{t} = \frac{G_{t}}{\sum(G_{t-s})* w_{s}}$ | **Equation 5:**  Take wastewater intensity or raw copies (G) and put it directly into the Rt equation for cases. |
| --- | --- |

##### 1.2.4 Rt method 4: Exponentiated change rate

At its core, the equation for the R_t_ by Cori et al. can be broken down into the change rate for incident cases during a time interval. We propose to calculate an R_t_ value that would be the change in wastewater detection since time t-1 to time t-0 to estimate the change rate in wastewater detection using the following equation:

| $R_{t}=e^{abs\left( \frac{G_{t-0}}{G_{t-1}-G_{t-0}} \right)}$ | **Equation 6:** Calculate the exponentiated change rate in raw gene copies over a specified time interval. |
| --- | --- |

The change rate in wastewater was centered around 1 and transforms wastewater concentration into a value akin to the R_t_.

##### 1.2.5 Rt method 5: Huisman method

Huismann et al. (12) analyzed the longitudinal data of raw wastewater influent of Zurich, Switzerland and primary sludge from San Jose, California, USA by combining these data with information on the shedding load distribution (SLD) to estimate a time series proportional to daily COVID-19 infection incidence. This approach provided robust estimates of R_t_ to compare with clinical data. Longitudinal data was first collected and combined with SLD. In the deconvolution process, the RNA information is profiled by SLD and normalization factor N, which estimates the measurement of viral RNA on day i related to past day incidence infection on day j. The study transformed RNA into a time series of infection incidence, then used the R package EpiEstim to estimate the R_t_ from incidence. While the method is independent of clinical case surveillance avoiding testing bias, the approach relies heavily on the SLD and the two normalization factors might be uncertain. Further, the method is susceptible to the variation in the observed data and “noisy” wastewater concentration data may lead to less accurate R_t_ estimates. For more information, see their GitHub: <https://github.com/JSHuisman/wastewaterRe>.

Huisman et al. (12) proposed a method for calculating the Rt from wastewater by first deconvolving wastewater data into approximate estimates of “cases” by dividing the detection levels of wastewater by the lowest detection level observed (purported to be akin to the baseline of one case detected). While the resulting case numbers are not on the same scale as actual incidence, the resulting values can then be used in the EpiEstim method for estimating Rt along with provided serial intervals for infection that Huisman et al. (12) derived from the literature. We adopt their method to compare with the other proposed methods here using the code that they published.

| $C_{i}=N*M\sum_{j} w_{i}-{{}_{j}I}_{j}$  $R_{t}= \frac{C_{i}}{\sum{(C_{i}}_{t-s})*w_{s}}$ | **Equation 7:** Gene copies are deconvolved into $C_{i}$ (a synthetic time series of case data) using the shedding load distribution of viral RNA from infections and two normalizing factors: $N$ for units of viral RNA per infection, and $M$ to account for variation between sewer systems. The synthetic case value $C_{i}$ is then used in the EpiEstim method to estimate $R_{t}$. |
| --- | --- |

##### 1.2.6 Rt method 6: Goldstein et al. method

Goldstein et al.(13) developed a compartmental model to estimate the reproductive number from concentrations of pathogen genomes in wastewater. That they call EIRR-ww model. The model has four compartments to produce the Rt estimate: E is the observed incidence, I is the unobserved incidence, R1 is the estimate recovered population, and R2 is the starting reproduction number based on case data. Goldstein et al. (13) propose the use of an EIRR model that is derived from and SEIR compartmental model structure without the S susceptible portion because they found the model to be more accurate. The model used a time-varying immigration rate( product of the proportion of susceptible individuals in the population and the transmission rate). They also tested a simulation and compared different similar models of case data and pathogen genomes to estimate the effective reproduction number of SARS-CoV-2. The model was also applied to estimate effective reproduction number of SARS-CoV-2 in Los Angeles, California, using pathogen RNA concentrations from wastewater. Goldstein et al. (13) also developed an SEIRR-ww model based on a standard compartmental EIRR model using a generalized t-distribution to model pathogen genome concentrations. We reproduce the EIRR-ww methods in the current paper because Goldstein et al. found that the EIRR-ww model was superior to the SEIRR-ww model.(13) The methods has several strengths including: avoiding assumptions about the dynamics of the susceptible population, pathogen genome concentrations are potentially less biased than case data, and the generalized t-distribution and accounts for the many outliers in wastewater data providing a good fit. There are also some weaknesses including that the model is complex involving several parameters to produce an inference and, while it avoids assumptions about the population, it still relies on parameters that might be accurately known. For more information, see their GitHub: <https://github.com/igoldsteinh/ww_paper>).

##### 1.2.7 Rt method 7: Episewer

The generative model proposed by Lison (14) contains five modules for infections, shedding, sewage, sampling, and measurements. The infections module employs a link function to ensure that Rt is always greater than 0 while still employing the smoothing approach selected. The smoothing approach selected is a smoothed spline that ensures a continuous time series of output. To model infections, the authors selected a stochastic renewal model (15). Either a Poisson or Negative binomial regression can be used to estimate realized infections. The shedding module uses an equation based on when expected symptom onset occurs to estimate the total load of virus shed into the sewer catchment. There are options for individual level variation, but generally, the programmer can match the observed cases for a period to the observed wastewater concentration along with flow to estimate the load-per-case. The sewage module is the continuously reported flow data. Next, the sampling module is the wastewater concentration data. This can be reported at any frequency. Last, the measurements module can make adjustments for factors like the limit of detection to tailor the approach to different concentration methods. All of these modules contribute to the final estimation method that is based on the observed wastewater concentration. Additional details are available from the EpiSewer GitHub repository and vignettes (<https://github.com/adrian-lison/EpiSewer/blob/main/vignettes/model-definition.md#user-content-fn-semimechanistic-6a82c85cb1e7bdfde831c52538434e6d>).

##### 1.2.8 Rt method 8: ERN

Champredon et al. (16) developed the effective reproduction number (ERN) package to estimate clinical and wastewater based effective reproduction numbers. For the wastewater-based approach, ERN takes a similar approach to that published by Huisman et al. (12) where the pathogen concentration is assumed to be a convolution of cases with a fecal shedding distribution proportional to the total pathogen shed by the population within the sewershed. ERN reports the following equation for estimating the concentration shed in wastewater:

| $w_{t}=\omega\sum_{k=1}^{i-1} i\left( t-k \right)f(k)$  $W_{t}=smooth interpolation(w_{t},\theta)$ | **Equation 8**: $\omega$ denotes the amount of viral concentration a single infection contributes. $W_{t}$ is the smoothed output from the individual, wastewater estimated case data. |
| --- | --- |

The signal from wastewater is then deconvolved using a smoothed interpolation of the wastewater data. ERN offers two smoothing approaches: the moving average or a LOESS. We used the LOESS in our application of the ERN package. Once daily incidence is obtained from the deconvolved wastewater data, ERN inputs those wastewater estimated cases into the EpiEstim model explained in equation 2.

### 2 Supplemental results

#### 2.1 EpiEstim – prior v. data-estimated serial interval

The serial interval (SI) for the EpiEstim method can either be provided or estimated from the data. We tested an SI of 4 with sd of 1 and the results were very similar to a data-estimated SI (Supplemental Figure 1). Thus, all of our models and comparisons that use the EpiEstim model for COVID-19 use an SI of 4 and sd of 1 to maintain efficient computation time. This had limited impact on final results.

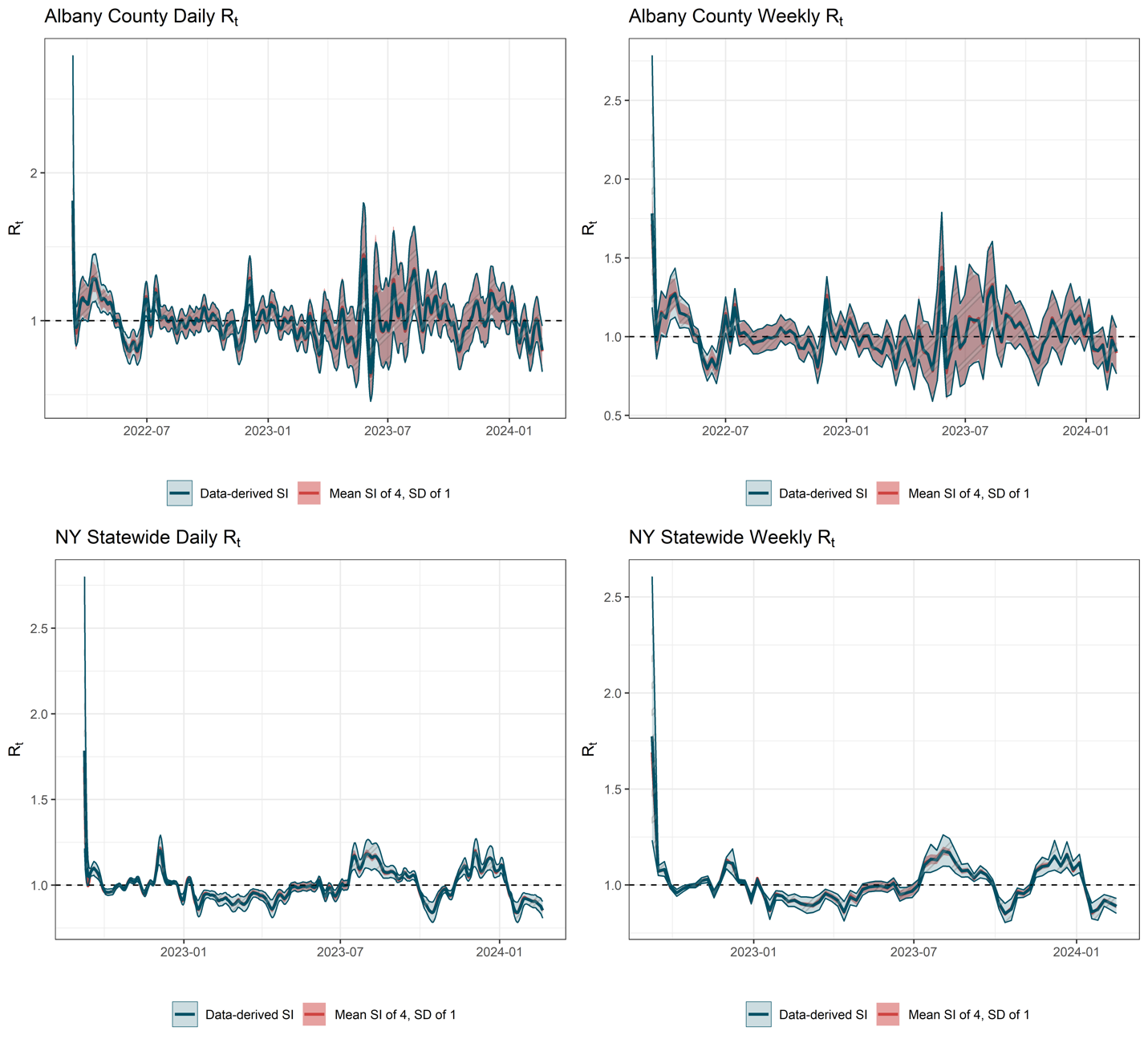

Supplemental Figure 1: EpiEstim Rt from case data with and without a known serial interval (SI).

#### 2.2 EpiEstim – different waves v. continuous

We wanted to ensure that Rt estimates across different waves would not be different if the estimations began at different times. We therefore compared the Rt estimates for a continuous time series to one that started later. The only observed difference was in the starting Rt model output. All other estimates after the initial estimate were the same (Supplemental Figure 2).

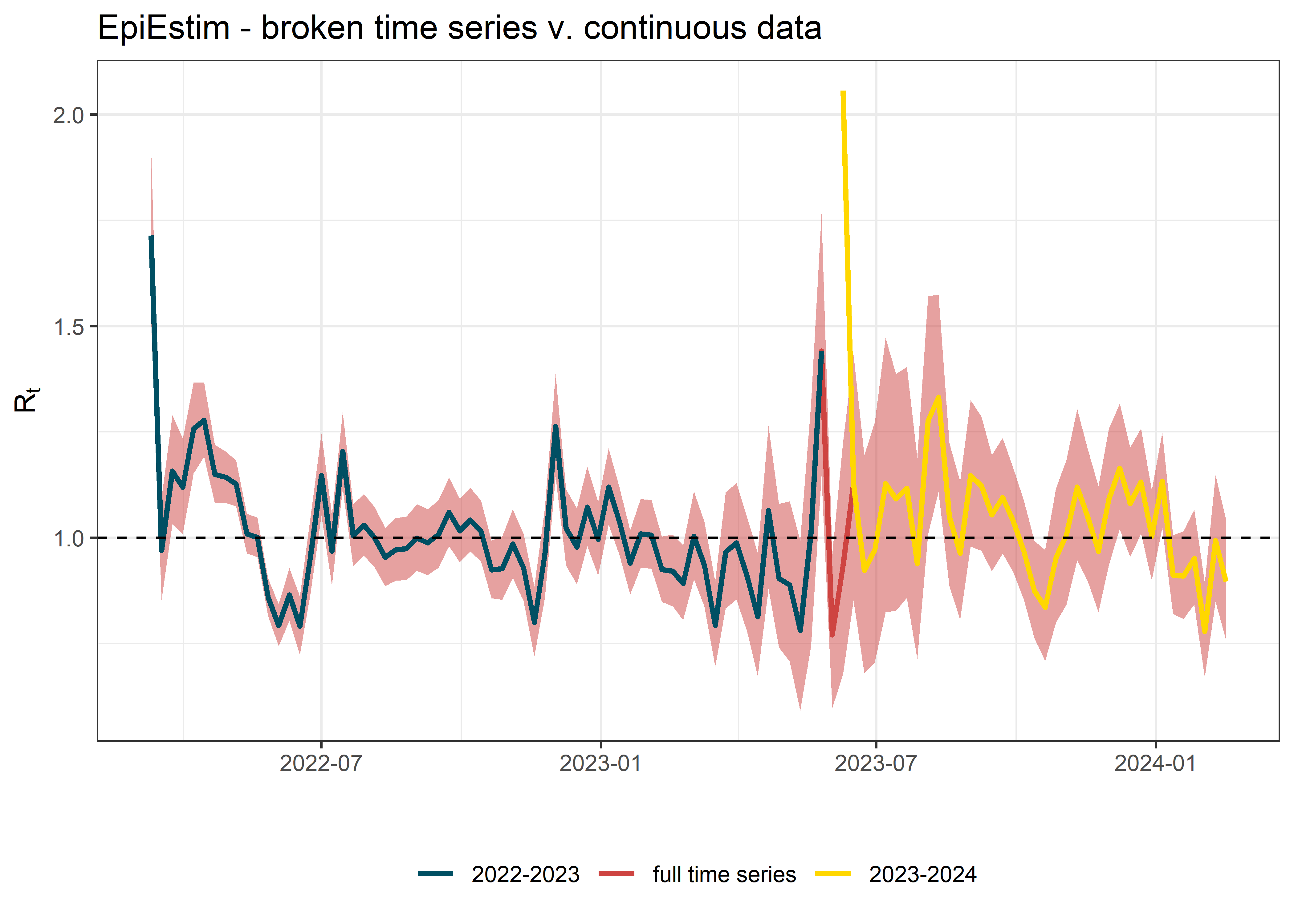

Supplemental Figure 2: A continuous time series estimate for the Rt as opposed to a broken time series split into two parts.

#### 2.3 Additional smoothing of wastewater data in models

Wastewater data tend to be more variable than case data even after smoothing the data using a 7-day average. Thus, we tested two additional rolling averages to see if wastewater data with less variability would produce Rt estimates closer to the case Rt estimates. We tested a 14-day average and a 30 day average. Based on comparing the Rt from wastewater for each of the averaging methods for the rolling GLM method to the Rt from case data, none of the smoothing methods was completely better (Supplemental Table ). While the 14-day average had higher percent of peaks that coincided, higher above or below 1 agreement, it also had equal sharpness to the 7-day average and worse RMSE. The 30 day average had the best Pearson correlation of 0.4447 followed by the 7-day average of 0.4161 and had lower mean absolute difference than the other two methods. Based on this, there are no definitive benefits to additionally smoothing of the wastewater data beyond the 7-day average. Additional smoothing can be used, however, without much measurable impact on the Rt estimation (Supplemental Table 2).

| Supplemental table 2: Evaluation metrics for the Rt from wastewater using the rolling GLM as compared to the Rt from case data for the entire state. | | | |
| --- | --- | --- | --- |
| *Metric* | *7-day smoothed average* | *14-day smoothed average* | *30-day smoothed average* |
| RMSE | 0.1360 | 0.1377 | 0.1413 |
| Pearson Correlation | 0.4161 | 0.4063 | 0.4447 |
| Percent of peaks that coincide | 0.1389 | 0.2581 | 0.1563 |
| Above or below 1 agreement | 0.6974 | 0.7763 | 0.7632 |
| Mean absolute difference | 0.0942 | 0.0916 | 0.0823 |
| Sharpness | 0.0010 | 0.0010 | 0.0011 |

#### 2.4 Additional smoothing of model outputs (Rt after model is run)

Model outputs for five of the methods were highly variable in their Rt estimates at the weekly time series aggregation. Thus we tested three rolling averages of the modeled output (3 weeks, 5 weeks, and 7 weeks). We found that at 5-week rolling average of the output from the wastewater Rt models best smoothed the results and complemented the case Rt better than the raw model outputs (Supplemental Figure 3).

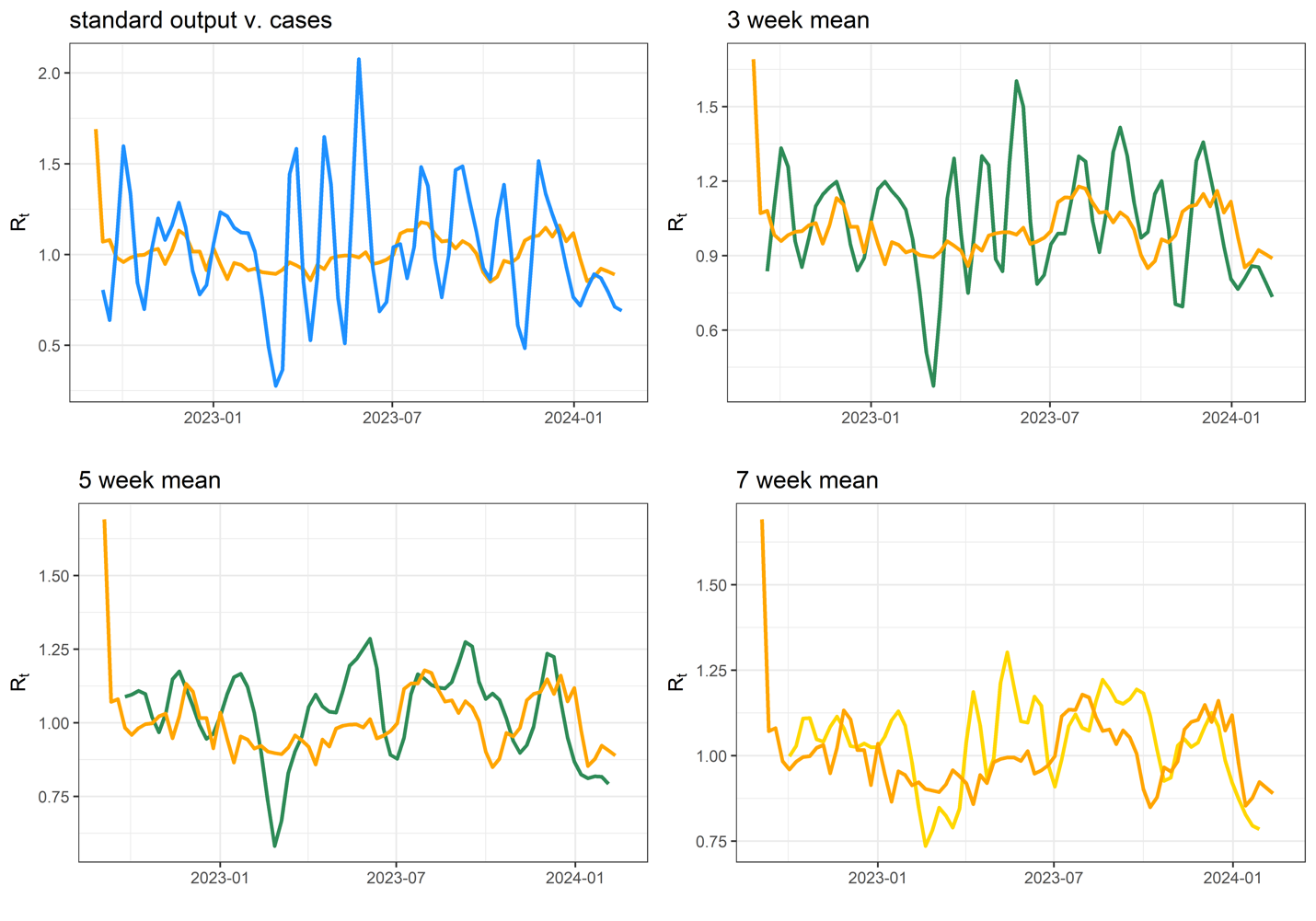

Supplemental Figure 3: Raw wastewater Rt output had higher variance than the output of the case Rt. To reduce variance, a 5 week rolling mean was selected to smooth the data and bring it more in alignment with the Rt from case data.

#### 2.5 Lag v. no lag

Wastewater concentrations for SARS-CoV-2 are known to precede increases in case reports by up to 4 days. We tested a daily lag of the wastewater data against new cases and did not find a lag to improve correlations (Supplemental Figure 4). Therefore, we ran all models without lagged data. Lagged and unlagged data did not vary in the final Rt estimates significantly (Supplemental Figure 5).

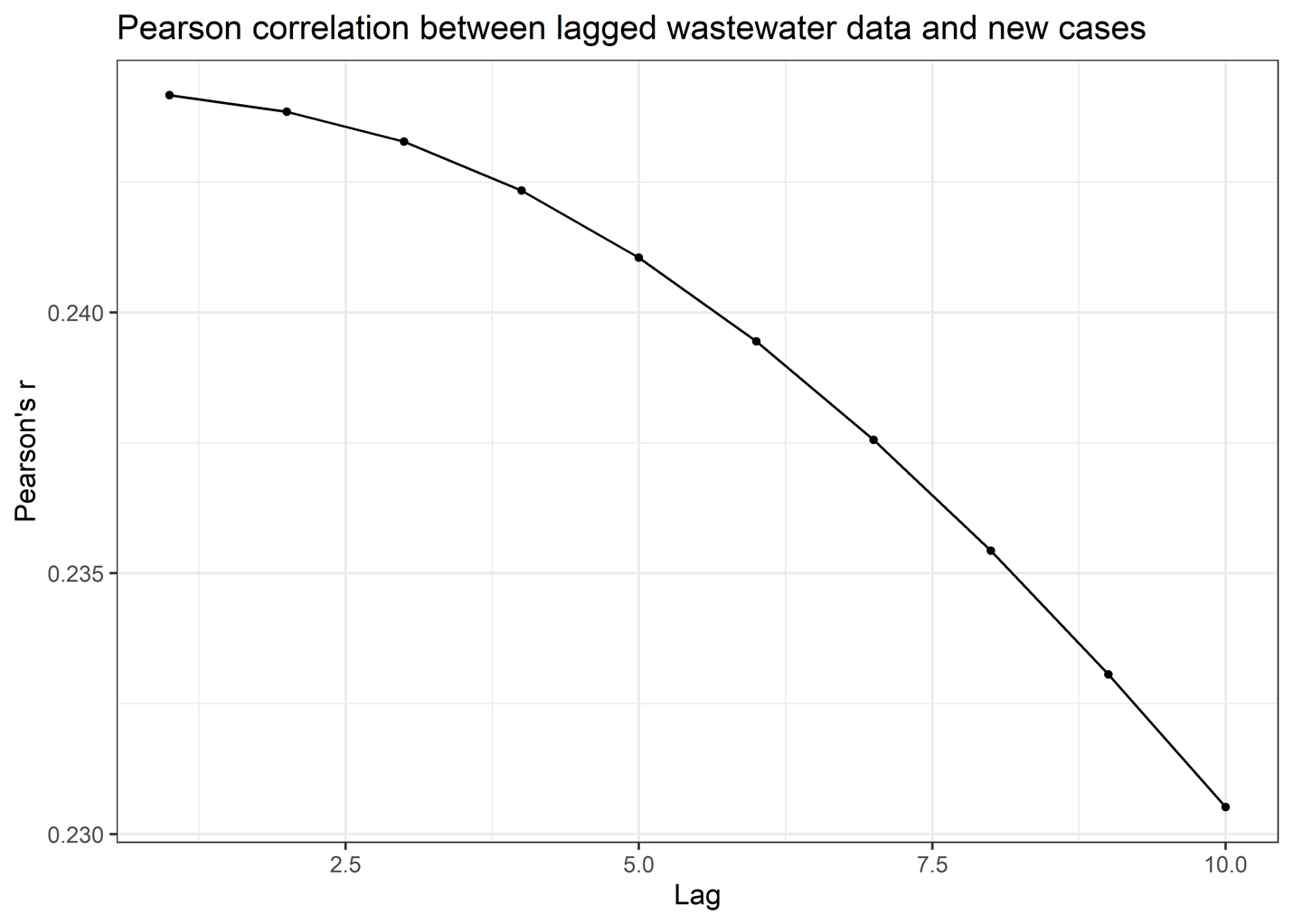

Supplemental Figure 4: The change in correlation between wastewater concentration and new case data over different time lags.

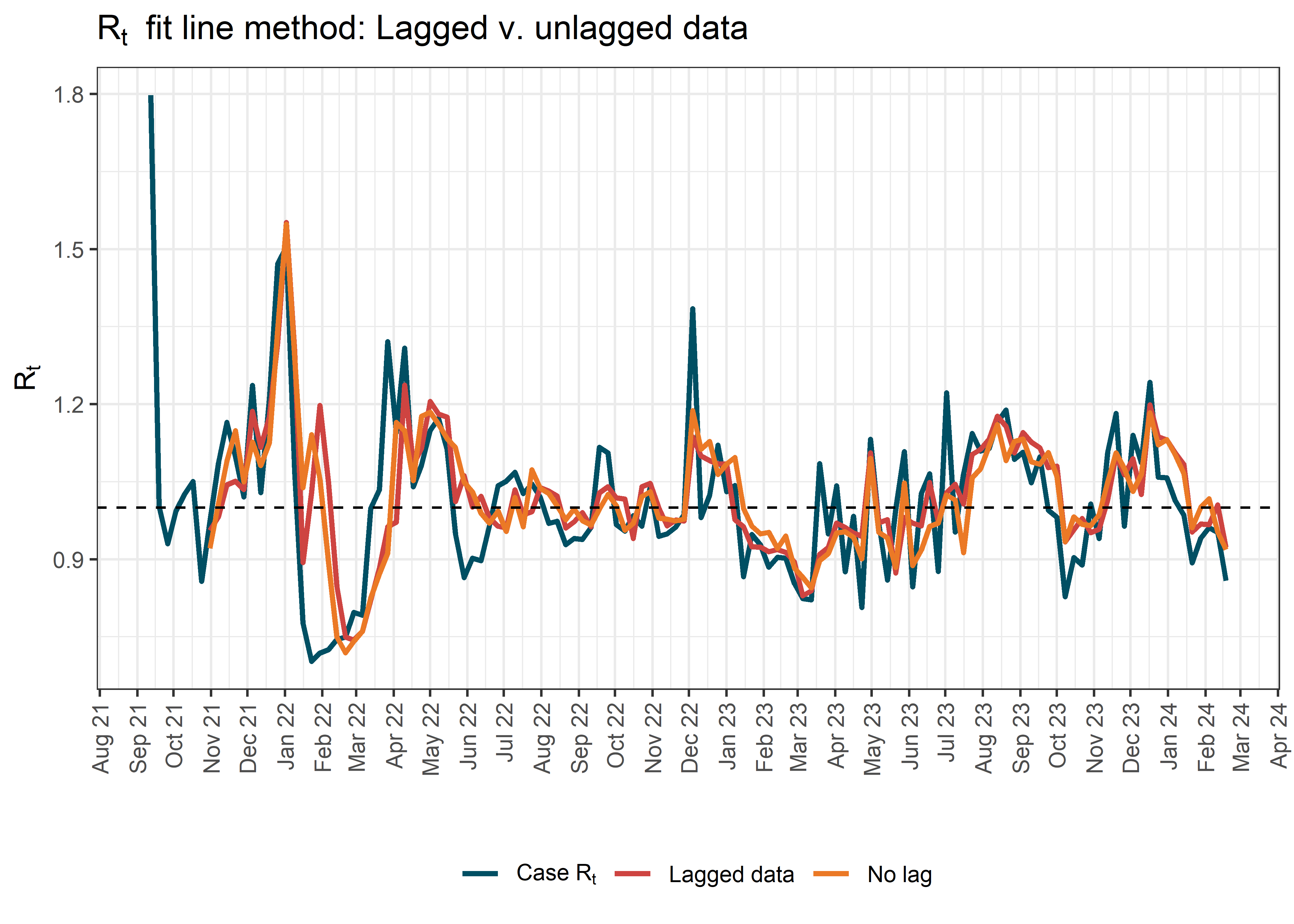

Supplemental Figure 5: Case Rt as compared to lagged wastewater Rt from the fit line method and unlagged data.

| Supplemental Table 3: Lagged data v. no lagged data for Rt calculation. There is a small difference in the values when the wasteater data are lagged 4 days. The unlagged data is closest to the reference model for the example county. | | | |
| --- | --- | --- | --- |
|  | *Fit line lagged* | *Fit line no lag* | *county* |
| RMSE | 0.106 | 0.106 | Orange |
| Pearson Correlation | 0.679 | 0.680 | Orange |
| Percent of peaks that coincide | 0.278 | 0.314 | Orange |
| Above or below 1 agreement | 0.688 | 0.711 | Orange |
| Mean absolute difference | 0.076 | 0.077 | Orange |
| Sharpness | 3.31 | 3.26 | Orange |

#### 2.6 Normalized v. non normalized

Normalization of wastewater data by a population factor like flow or human fecal indicator is a common practice. We tested the Rt methods with population normalized data and the estimates did not change dramatically (Supplemental Figure 6). Therefore, given that we did not have flow or fecal indicator data for all sites, we determined to use the raw data in our analysis. Also, many methods like EpiSewer are optimal with raw data.

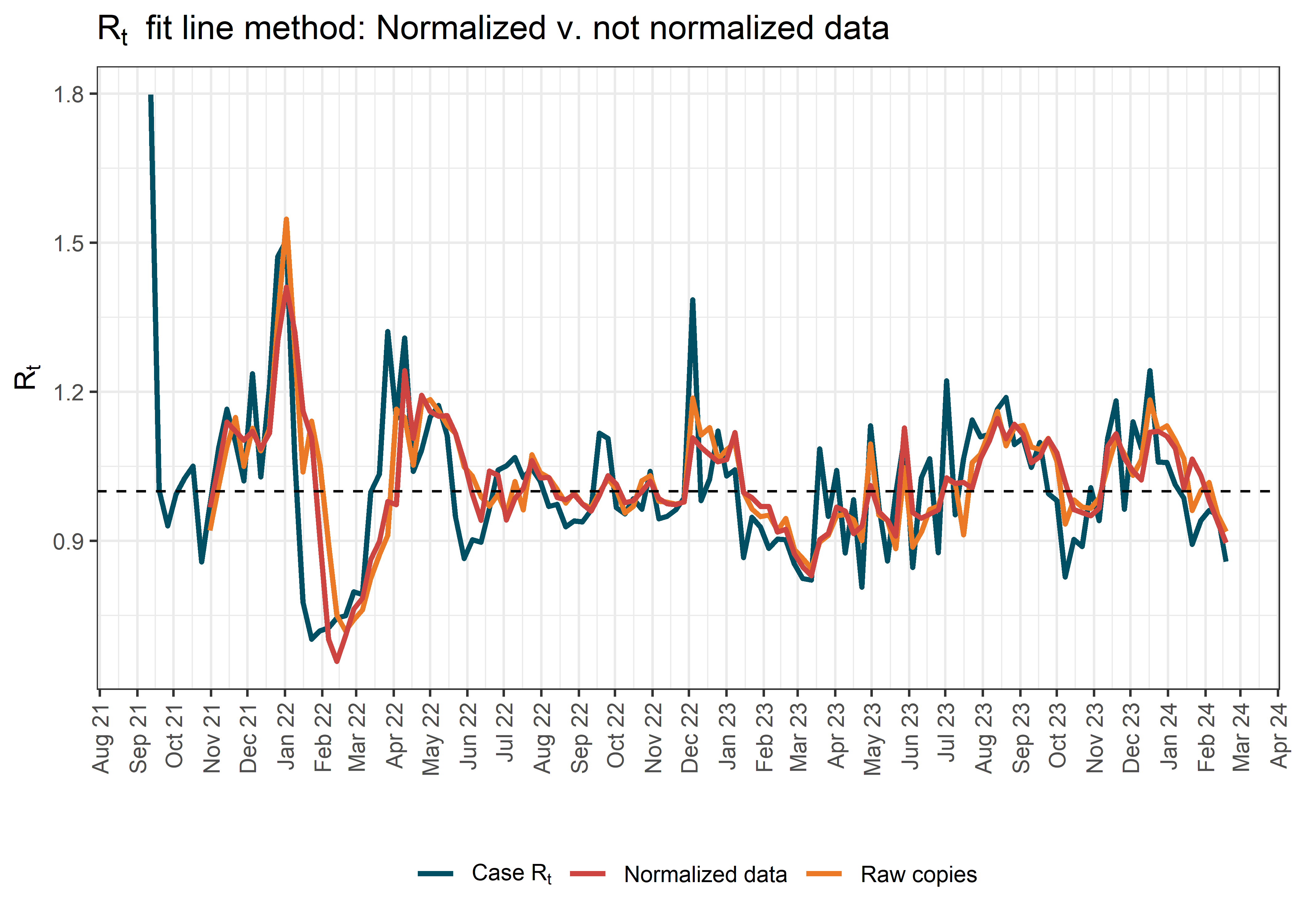

Supplemental Figure 6: Normalized wastewater Rt estimates and Rt estimates from raw data.

| Supplemental Table 4: Normalized data v. not normalized data for Rt calculation. There is a small difference in the values when the wasteater data are lagged 4 days. Neither method is necessarily superior. | | | |
| --- | --- | --- | --- |
|  | *Fit line raw copies* | *Fit line normalized* | *county* |
| RMSE | 0.106 | 0.106 | Orange |
| Pearson Correlation | 0.680 | 0.677 | Orange |
| Percent of peaks that coincide | 0.314 | 0.294 | Orange |
| Above or below 1 agreement | 0.711 | 0.734 | Orange |
| Mean absolute difference | 0.077 | 0.076 | Orange |
| Sharpness | 3.26 | 5.66 | Orange |

#### 2.7 With or without New York City

New York State has ten primary regions with one being New York City (NYC). Given the high population density of NYC, it sometimes biases models toward estimates that only reflect the city’s condition. Therefore, we fit a model with and without data from NYC to determine the impact on the statewide case Rt. We found that there was little impact on the Rt estimates and therefore, we determined to keep NYC in all models (Supplemental Figure 7).

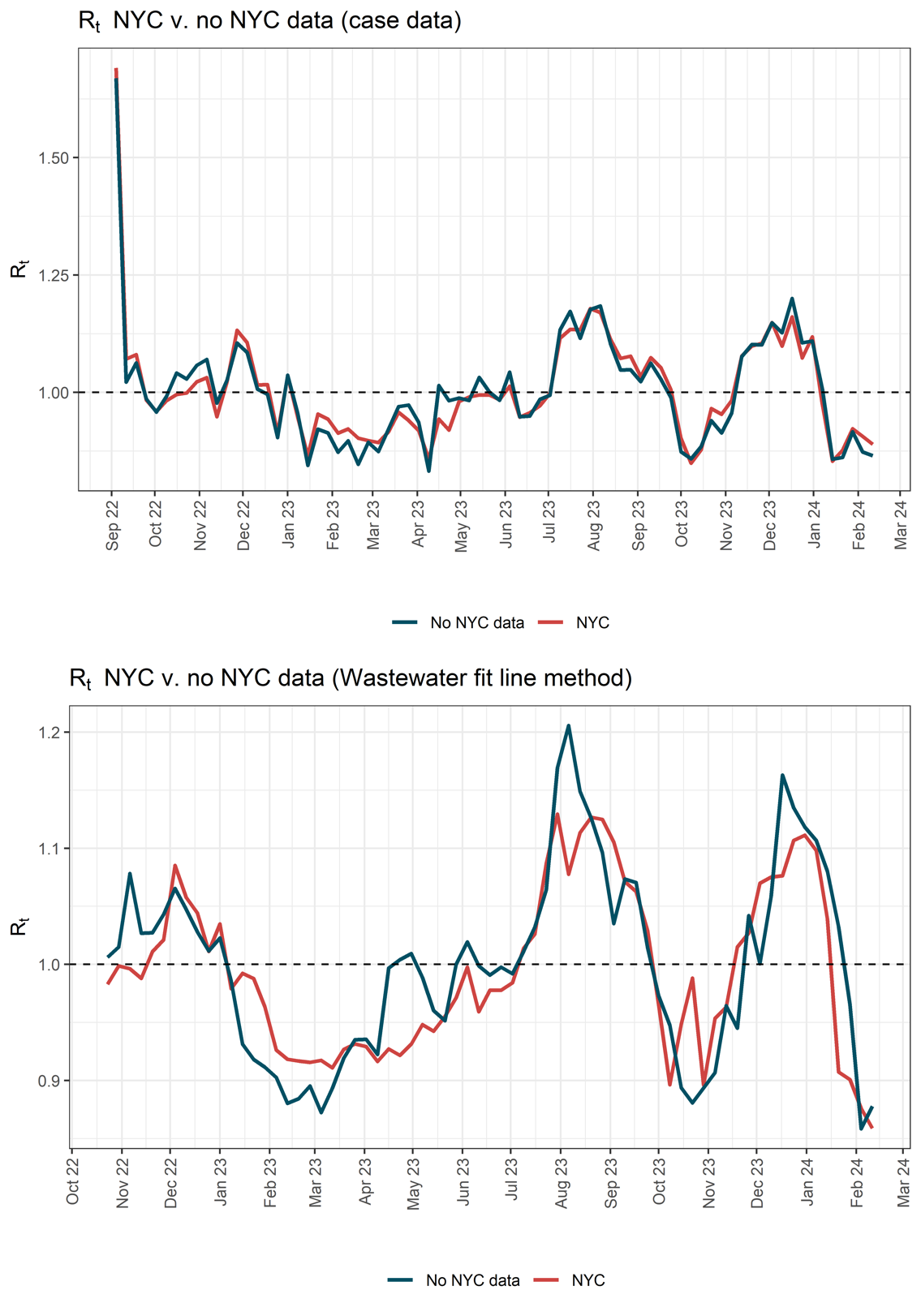

Supplemental Figure 7: Rt estimates with data for all of NYS and estimates from models that excluded data fro mNYC

#### 2.8 ERN exploration

The ERN model has some additional parameters that we tested to help select the optimal situation for NYS. We selected a LOESS smooth and a scaling factor of 0.7. We also tested if the initial Omicron surge adversely impacted estimates of the entire time period. We determined that the length of the data also was linked to the need for different span widths for the LOESS smooth. We determined that data with less than 200 observations produced optimal estimates with a LOESS of 0.2, data with 200 to 500 observations produced optimal estimates with a LOESS of 0.15, and data with more than 500 observations produced an optimal estimate with a LOESS span of 0.1. See Supplemental Figure 8 for different scenarios we tested.

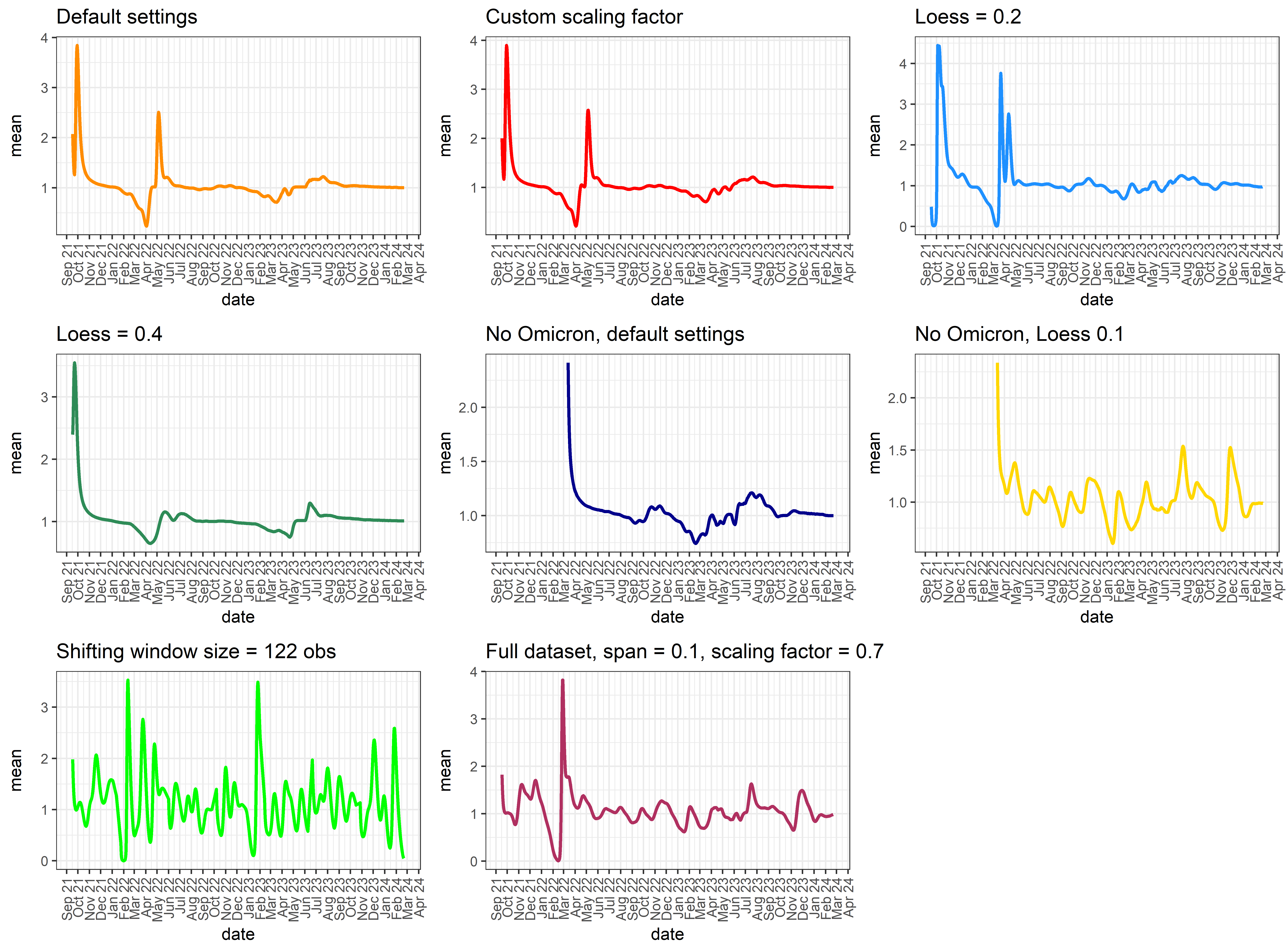

Supplemental Figure 8: Different model fitting situations for ERN.

#### 2.9 Log v. not logged transformation

Log transformation of wastewater data is often done to improve the linear fit of the concentrations to match with clinical data. We found that log transformation prior to model fitting was not necessary and actually hindered model performance (Supplemental Figure 9).

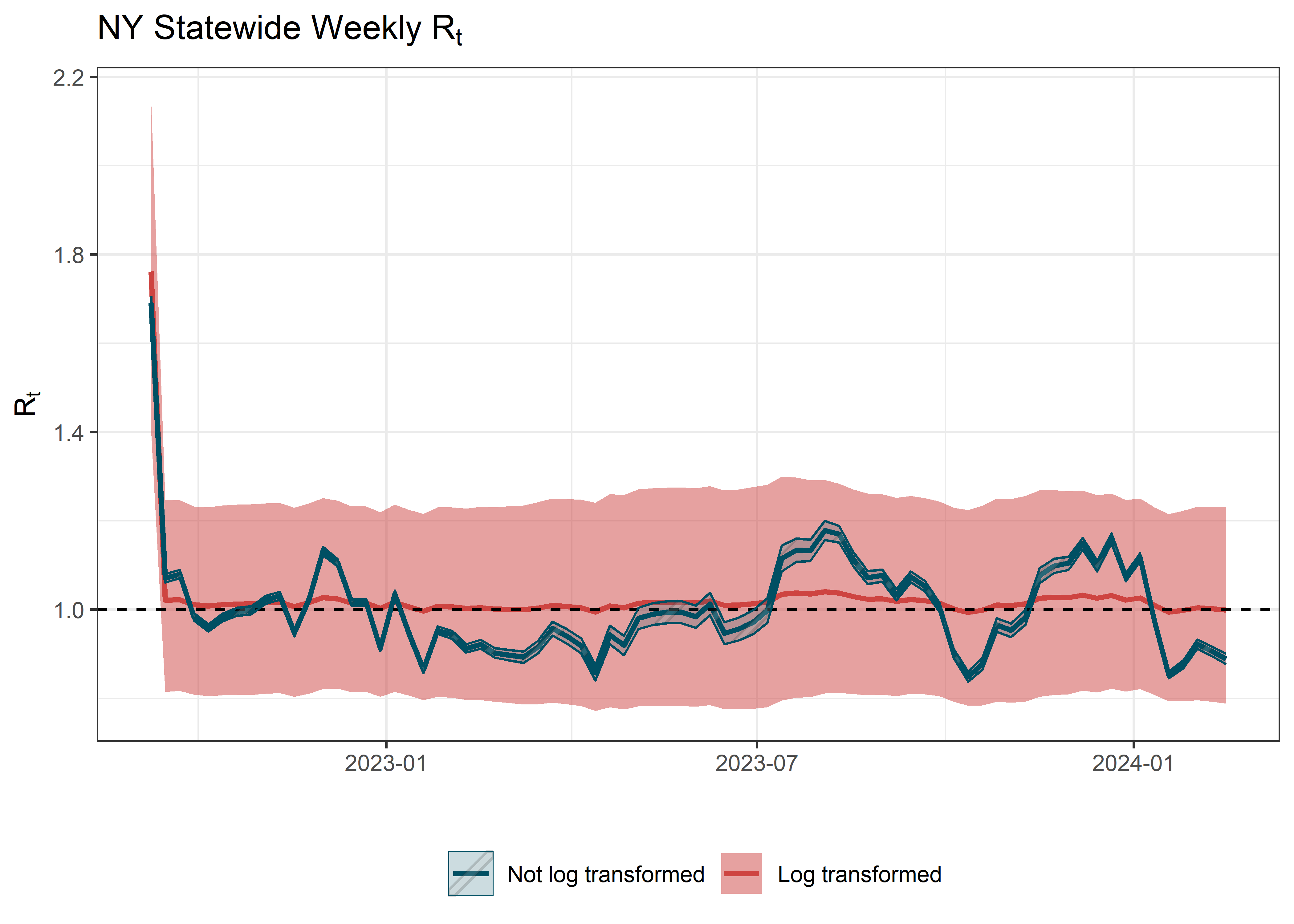

Supplemental Figure 9: Rt from wastewater with and without log-transformed data.

### 3 References

1.	Hill DT, Alazawi MA, Moran EJ, Bennett LJ, Bradley I, Collins MB, et al. Wastewater surveillance provides 10-days forecasting of COVID-19 hospitalizations superior to cases and test positivity: A prediction study. Infect Dis Model. 2023 Dec 1;8(4):1138–50.

2.	Hoar C, Chauvin F, Clare A, McGibbon H, Castro E, Patinella S, et al. Monitoring SARS-CoV-2 in wastewater during New York City’s second wave of COVID-19: sewershed-level trends and relationships to publicly available clinical testing data. Environ Sci Water Res Technol. 2022 May 5;8(5):1021–35.

3.	Wilder ML, Middleton F, Larsen DA, Du Q, Fenty A, Zeng T, et al. Co-quantification of crAssphage increases confidence in wastewater-based epidemiology for SARS-CoV-2 in low prevalence areas. Water Res X. 2021 May 1;11:100100.

4.	Lu X, Wang L, Sakthivel SK, Whitaker B, Murray J, Kamili S, et al. US CDC Real-Time Reverse Transcription PCR Panel for Detection of Severe Acute Respiratory Syndrome Coronavirus 2. Emerg Infect Dis. 2020 Aug;26(8):1654–65.

5.	Cori A, Ferguson NM, Fraser C, Cauchemez S. A New Framework and Software to Estimate Time-Varying Reproduction Numbers During Epidemics. Am J Epidemiol. 2013 Nov 1;178(9):1505–12.

6.	Cori A. EpiEstim: Estimate Time Varying Reproduction Numbers from Epidemic Curves [Internet]. 2021. Available from: https://CRAN.R-project.org/package=EpiEstim

7.	Gostic KM, McGough L, Baskerville EB, Abbott S, Joshi K, Tedijanto C, et al. Practical considerations for measuring the effective reproductive number, Rt. PLOS Comput Biol. 2020 Dec 10;16(12):e1008409.

8.	Larsen DA, Collins MB, Du Q, Hill D, Insaf TZ, Kilaru P, et al. Coupling freedom from disease principles and early warning from wastewater surveillance to improve health security. PNAS Nexus. 2022 Mar 1;1(1):pgac001.

9.	Prasek SM, Pepper IL, Innes GK, Slinski S, Betancourt WQ, Foster AR, et al. Variant-specific SARS-CoV-2 shedding rates in wastewater. Sci Total Environ. 2023 Jan 20;857:159165.

10.	Montesinos-López JC, Daza-Torres ML, García YE, Herrera C, Bess CW, Bischel HN, et al. Bayesian sequential approach to monitor COVID-19 variants through test positivity rate from wastewater. mSystems. 2023 Jul 25;8(4):e00018-23.

11.	Daza-Torres ML, Montesinos-López JC, Kim M, Olson R, Bess CW, Rueda L, et al. Model training periods impact estimation of COVID-19 incidence from wastewater viral loads. Sci Total Environ. 2023 Feb 1;858:159680.

12.	Huisman JS, Scire J, Caduff L, Fernandez -Cassi Xavier, Ganesanandamoorthy P, Kull A, et al. Wastewater-Based Estimation of the Effective Reproductive Number of SARS-CoV-2. Environ Health Perspect. 2022;130(5):057011.

13.	Goldstein IH, Parker DM, Jiang S, Minin VM. Semiparametric inference of effective reproduction number dynamics from wastewater pathogen surveillance data. ArXiv. 2023 Aug 31;arXiv:2308.15770v2.

14.	Lison A. adrian-lison/EpiSewer [Internet]. 2024 [cited 2024 Aug 22]. Available from: https://github.com/adrian-lison/EpiSewer

15.	Bhatt S, Ferguson N, Flaxman S, Gandy A, Mishra S, Scott JA. Semi-mechanistic Bayesian modelling of COVID-19 with renewal processes. J R Stat Soc Ser A Stat Soc. 2023 Oct 1;186(4):601–15.

16.	Champredon D, Papst I, Yusuf W. ern: An R package to estimate the effective reproduction number using clinical and wastewater surveillance data. PLOS ONE. 2024 Jun 21;19(6):e0305550.
