## Supplementary figures and images for "Estimating the effective reproduction number from wastewater (R_t_): A methods comparison"

### decision tree

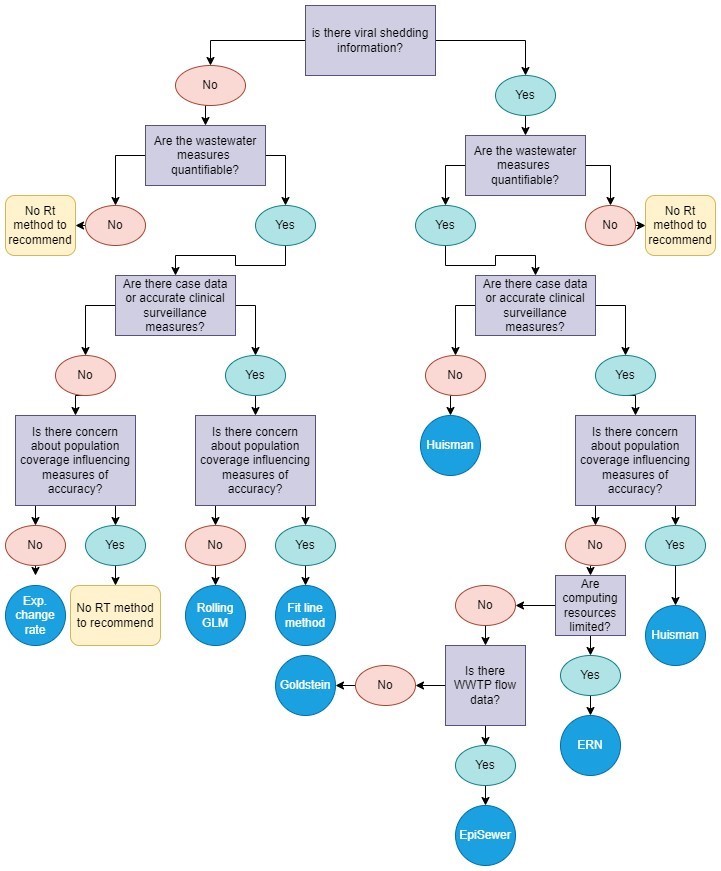
